# A genotype-phenotype taxonomy of hypertrophic cardiomyopathy

**DOI:** 10.1101/2023.03.11.23285908

**Authors:** Lara Curran, Antonio de Marvao, Paolo Inglese, Kathryn A McGurk, Pierre-Raphaël Schiratti, Adam Clement, Sean L Zheng, Surui Li, Chee Jian Pua, Mit Shah, Mina Jafari, Pantazis Theotokis, Rachel J Buchan, Sean J Jurgens, Claire E Raphael, Arun John Baksi, Antonis Pantazis, Brian P Halliday, Dudley J Pennell, Wenjia Bai, Calvin W L Chin, Rafik Tadros, Connie R Bezzina, Hugh Watkins, Stuart A Cook, Sanjay K Prasad, James S Ware, Declan P O’Regan

## Abstract

**Background:** Hypertrophic cardiomyopathy (HCM) is an important cause of sudden cardiac death associated with heterogeneous phenotypes but there is no systematic framework for classifying morphology or assessing associated risks. Here we quantitatively survey genotype-phenotype associations in HCM to derive a data-driven taxonomy of disease expression.

**Methods:** We enrolled 436 HCM patients (median age 60 years; 28.8% women) with clinical, genetic and imaging data. An independent cohort of 60 HCM patients from Singapore (median age 59 years; 11% women) and a reference population from UK Biobank (n = 16,691, mean age 55 years; 52.5% women) were also recruited. We used machine learning to analyse the three-dimensional structure of the left ventricle from cardiac magnetic resonance imaging and build a tree-based classification of HCM phenotypes. Genotype and mortality risk distributions were projected on the tree.

**Results:** Carriers of pathogenic or likely pathogenic variants for HCM (P/LP) variants had lower left ventricular mass, but greater basal septal hypertrophy, with reduced lifespan (mean follow-up 9.9 years) compared to genotype negative individuals (hazard ratio: 2.66; 95% confidence interval [CI]: 1.42-4.96; *P* < 0.002). Four main phenotypic branches were identified using unsupervised learning of three-dimensional shape: 1) non-sarcomeric hypertrophy with co-existing hypertension; 2) diffuse and basal asymmetric hypertrophy associated with outflow tract obstruction; 3) isolated basal hypertrophy; 4) milder non-obstructive hypertrophy enriched for familial sarcomeric HCM (odds ratio for P/LP variants: 2.18 [95% CI: 1.93-2.28, *P* = 0.0001]). Polygenic risk for HCM was also associated with different patterns and degrees of disease expression. The model was generalisable to an independent cohort (trustworthiness *M*_*1*_: 0.86-0.88).

**Conclusions:** We report a data-driven taxonomy of HCM for identifying groups of patients with similar morphology while preserving a continuum of disease severity, genetic risk and outcomes. This approach will be of value in understanding the causes and consequences of disease diversity.

## Introduction

Hypertrophic cardiomyopathy (HCM) is an inherited cardiac condition (prevalence ∼1 in 500) related to increased risk of sudden death and adverse cardiac events, including in early life and middle age, which is associated with genetic and phenotypic heterogeneity.^1^ Although traditionally considered a Mendelian disease, polygenic variation is now recognised as contributing to phenotypic variability in carriers of HCM-causing rare sarcomeric variants.^2,3^ Sex and environmental risk factors also interact with disease-associated variants to modify susceptibility.^4^ HCM-associated rare variants are not infrequently observed in the general population, but the most prevalent variants cause an attenuated phenotype and lower risk of adverse events outside the context of familial disease.^5^ In the more common non-sarcomeric HCM there is substantial polygenic inheritance and modifiable risk factors have important roles in disease expressivity.^3^ HCM is also not a static condition and adverse remodelling and fibrosis can evolve over time.^6^ Such dynamic endophenotypic diversity presents challenges for understanding drivers of heterogeneity, identifying patients enriched for pathogenic variants, and for developing personalized clinical profiles to guide intervention.

Current approaches to improve patient stratification in HCM, using sarcomere variant status and morphological traits, have described potential functional and anatomic groupings with differing outcomes.^7,8^ However, such approaches do not align with the molecular understanding of HCM as a continuum of phenotypic expression influenced by genetic and environmental modifiers.^9^ While optimal care requires cardiac imaging to confirm a diagnosis of HCM and characterize individual pathophysiology,^10^ there is limited understanding of phenotypic diversity and its relevance to genotype status and clinical management. Here we apply novel approaches for phenotyping patients using quantitative three dimensional representations of morphology to map the genotype-phenotype architecture of HCM and to discover a “taxonomy” that identifies similar groups of patients while preserving a continuous distribution of risk. We propose this as a data-driven framework for visualizing individual patient profiles in relation to morphological phenotype across a spectrum of sarcomeric and non-sarcomeric HCM.

## Methods & Materials

### Overview

We applied computational techniques to create patient-specific 3D models of the left ventricle from cardiac magnetic resonance imaging (CMR) (Fig. 1a). We used these to create a regional model of wall thickness and geometry in genotyped patients with HCM and in a control population from UK Biobank (UKB) to visualise aggregated genotype-phenotype associations. We applied discriminative dimensionality reduction to build a tree-like classification of HCM where phenotypes and risks are continuously distributed while groups of morphologically similar patients are clustered together. We tested the validity of our approach by mapping an external cohort of HCM patients of different ancestry to our tree structure and assessed the similarity of phenotypic distribution between these groups.

**Figure 1.**
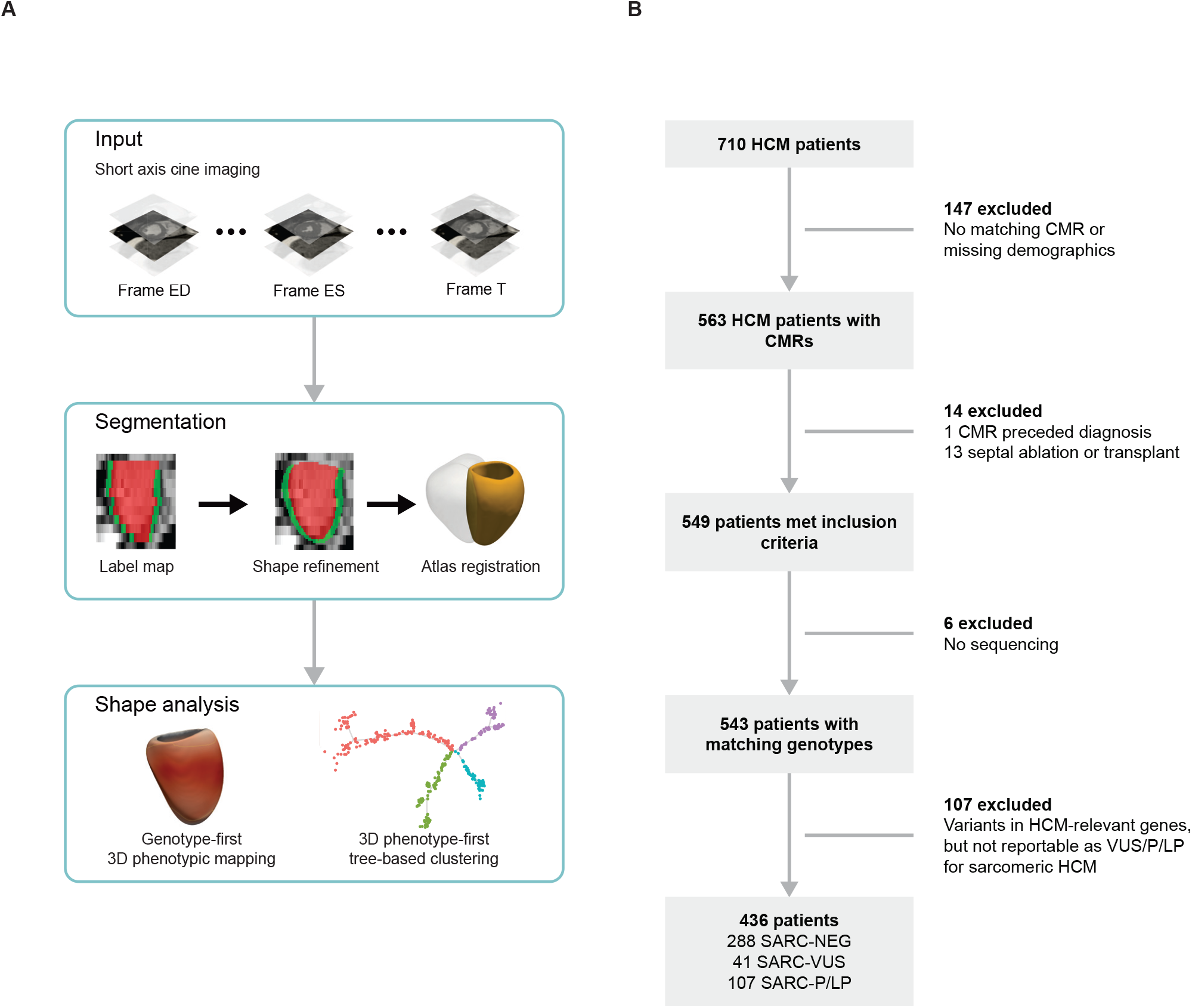
Study flowchart. **a**. Details of the analysis pipeline using segmentations of cardiac magnetic resonance cine imaging to build a 3D model of phenotypic variation in UK Biobank and HCM participants. **b**. Details of patients with hypertrophic cardiomyopathy (HCM) recruited to the study and reasons for exclusion. CMR, cardiac magnetic resonance; ED, end diastole; ES, end systole; SARC-NEG, those without variants in genes that may cause or mimic HCM; SARC-P/LP, pathogenic or likely pathogenic sarcomeric variants; SARC-VUS, variants of uncertain significance; T, last cardiac phase.

### HCM participants

In total 710 patients with a clinical diagnosis of HCM, either seen in the inherited cardiomyopathy service or referred for CMR imaging, were consecutively enrolled into a prospective registry at the National Institute for Health Research (NIHR) Royal Brompton Hospital Cardiovascular Biobank project between 2009-2015, of whom 436 were included in this study. All participants provided written informed consent and the study was approved by the National Research Ethics Service (19/SC/0257). HCM diagnosis was independently adjudicated by a cardiomyopathy specialist based on established clinical and CMR criteria where all patients met the American Heart Association criteria for diagnosis.^10^ This was defined as a wall thickness of 15mm or greater, or 13–14mm if there was a first degree relative with HCM, not explained by another cardiac or systemic disease-causing abnormal loading conditions, or had disproportionate apical wall thickness and tapering in keeping with an apical HCM phenotype.^11^

Patients were excluded from analysis based on age (< 16 years at time of CMR), missing demographic or clinical data, contraindication to CMR, previous history of septal ablation, cardiac transplantation or myectomy at baseline (Fig. 1b). A history of hypertension or diabetes was documented, as well as current medication at time of enrolment to the study. The cohort underwent detailed clinical, imaging and genetic assessment. All patients underwent CMR for assessment of cardiac chamber volumes and function (1.5T, Siemens Sonata or Avanto, Siemens Medical Systems, Erlangen, Germany). Variables reported were collected at enrolment to the study. The CMRs selected for analysis were those closest to the date of enrolment or the first diagnostic study available. Where present, left ventricular outflow tract obstruction (LVOTO) was confirmed through stress echocardiography. Unrelated Singaporean patients with a diagnosis of HCM (n = 60) were prospectively recruited from the National Heart Centre Singapore. Patients gave written informed consent to participate which was approved by the Singhealth Centralised Institutional Review Board (2020/2353) and Singhealth Biobank Research Scientific Advisory Executive Committee (SBRSA 2019/001v1). Singaporean subjects underwent an equivalent CMR protocol at 1.5T (Aera, Siemens, Erlangen, Germany) or 3T (Ingenia, Philips, Best, Netherlands).

Conventional CMR analysis was undertaken by accredited operators using semi-automated software (CMRtools, Cardiovascular Imaging Solutions, London, UK).

### UK Biobank participants

The UKB study recruited 500,000 participants aged 40 to 69 years old from across the United Kingdom between 2006 and 2010 (National Research Ethics Service, 11/NW/0382).^12^ This study was conducted under terms of access approval number 40616. In each case, written informed consent was provided.

A sub-study of UKB invited participants for CMR for assessment of cardiac chamber volumes and function using a standard protocol (1.5T, Siemens Aera, Siemens Medical Systems, Erlangen, Germany).^13^ As a reference population, we selected 16,691 participants that did not meet criteria for left ventricular hypertrophy and were classified as genotype negative (SARC-NEG) by having no variants in genes that may cause or mimic HCM (see Sequencing and variant categorisation).

### Cardiac phenotyping using machine learning

Segmentation of the cine images in both UKB and HCM groups was performed using a deep learning neural network algorithm developed and optimised in-house. The performance of image annotation using this algorithm is equivalent to a consensus of expert human readers and achieves sub-pixel accuracy for cardiac segmentation.^14^ The label maps were super-resolved and registered to a cardiac atlas enabling consistent quantitative three-dimensional phenotypic analysis within and between patient groups.^15^

Myocardial wall thickness was measured along radial line segments connecting the endocardial and epicardial surfaces perpendicular to the myocardial centreline and excluding trabeculae. Chamber volumes and mass were calculated from the segmentations according to standard post-processing guidelines.^16^ Myocardial strain analysis was performed using non-rigid free-form deformation image registration.^17^ Trabecular traits were quantified using fractal dimension (FD) analysis where a higher value indicates more complex trabeculation.^18^

### Sequencing and variant categorisation

Panel sequencing was completed in the HCM patient cohort, as previously described.^19^ The patients were sequenced using either a custom SureSelect capture panel targeting genes associated with inherited cardiac conditions or the Illumina TruSight Cardio panel. Sequencing was performed on either the SOLiD 5500xl platform, or the Illumina HiSeq, MiSeq or NextSeq platforms. The Singaporean cohort underwent targeted genetic sequencing using the TruSight Cardio panel and an equivalent pipeline as previously reported.^20^

Patients were divided into three genetic strata. Patients carrying at least one potentially-causative rare variant (allele frequency <0.00004)^21^ in any of 8 sarcomere-encoding genes robustly associated with HCM were considered genotype positive. These were further stratified into (i) those carrying variants previously confidently classified as pathogenic / likely pathogenic (SARC-P/LP) in ClinVar, and confirmed on our review, or else curated as P/LP according to ACMG criteria using the semi-automated CardioClassifier decision support tool^22^ (n = 107), as previously published,^5^; and (ii) those carrying sarcomeric variants of uncertain significance (SARC-VUS), comprising variants in the same 8 genes, that are consistent with known disease mechanisms and sufficiently rare, but with insufficient evidence to classify robustly as P/LP.^23,24^

Individuals were classified as genotype negative (SARC-NEG) if they had no rare protein-altering variant (minor allele frequency <0.001 in the UKB and the Genome Aggregation Database)^25^ in any of 25 genes that potentially cause HCM (definitive or moderate evidence according to international curation)^26^ or cause syndromes that can present with isolated left ventricular hypertrophy (genocopies).^26^ In order to generate the most robust set of true genotype negatives, individuals carrying a protein-altering variant in any these 25 genes, but that was not sufficiently rare to be considered potentially causative of monogenic HCM was excluded from the analysis. Further details are given in Supplementary Materials. Common genetic variation contributes substantially to HCM risk and we also assessed the relationship between phenotype and polygenic score (PGS) derived from a case-control HCM genome wide association study (GWAS) in the 100,000 Genomes Project.^27^

### Outcome measures

Data were collected to measure all-cause mortality in the HCM cohort. Outcomes were verified through search of the NHS Shared Care Records. Patients were followed up for a median of 10.2 years from date of study enrollment.

### Statistical analysis and data modelling

Statistical analysis was performed with R (version 4.0.3) and RStudio Server (version 1.2; Boston, MA), unless otherwise stated. Variables were expressed as percentages if categorical, mean ± standard deviation (SD) if continuous and normal, and median ± inter-quartile range (IQR) if continuous and non-normal. Baseline anthropometric data were compared by Kruskal-Wallis tests and, if differences were identified, a Wilcoxon test was used for pairwise comparisons with BenjaminiHochberg adjustment for multiple testing. Clustering of clinical data from HCM patients was performed using UMAP (uniform manifold approximation and projection).^28^

The association between genotype and three-dimensional phenotype was assessed using vertex-wise regression modeling, controlling for false discovery, plotting the strength of association between regional wall thickness and shape on the epicardial surface of the models.^5,29^ Clustering of subjects by their 3D left ventricular wall thickness, adjusted for age, sex and ancestry, was performed through partitioning the shared nearest neighbour (SNN) graph^30^ with the multilevel refinement Leiden algorithm.^31^ The SNN graph and its partitions were determined using the functions available in Seurat.^32^ The clusters were visually inspected by UMAP projection and their stability was assessed through bootstrapping using fpc.^33^ We used DDRTree (discriminative dimensionality reduction via learning a tree) to project the 3D left ventricular wall thickness into a 2D tree structure to visualise the distribution of HCM phenotypes.^34,35^

The predictive power of the DDRTree mapping for P/LP genotype was tested with a generalized additive model (GAM) fitted on the tree coordinates, using a 10-fold cross validation repeated 3 times. Survival probability to median observed age was estimated with a Cox proportional hazards model fitted on the cubic spline of tree coordinates of each individual and their relative position in the tree. Association between the DDRTree mapping and polygenic risk score was assessed with a logistic regression GAM model using the subjects’ coordinates as independent variables and the binarized polygenic risk score (thresholded at median) as outcome. The predictions were obtained using a 10-fold cross validation.

The primary survival analysis was performed in individuals from the HCM cohort with chronological age as time-scale and adjusting for genetic sex and ancestry (dichotomised by white European ancestry). Participants with SARC-P/LP variants were compared with pooled participants without variants and with SARC-VUS carriers. Proportional hazards assumption as assessed using Schoenfeld residuals was not violated.

## Results

### Participants

The HCM cohort consisted of 436 eligible patients of whom 287 (66.0%) were classified as SARC-NEG, 41 (9.4%) as SARCVUS and 107 (24.6%) as SARC-P/LP. Most were European (n = 352; 80.1%) and men (n = 310; 71.1%). Patient demographics and CMR-derived measurements are stratified by genotype in Table 1. There were 16,691 UKB participants (Supplemental Table 5) selected for the absence of HCM, and the absence of any rare variant in a gene associated with HCM or a potential genocopy (European = 14,683, 87.9%; women 52.5%, age 55 ± 7.5 years). Of the 60 Singaporean HCM patients (Chinese = 52, 86.7%; women 11.7%; median age 58.9, IQR: 46-66), 28 (46.6%) were classified as SARC-NEG, 16 (26.7%) as SARC-VUS and 16 (26.7%) as SARC-P/LP (Supplemental Table 6).

**Table 1.** Patient characteristics and CMR-derived measurements by genotype. Mean ± standard deviation. *P* value only shown if nominally significant in at least one comparison. BSA, body surface area; concentricity, (left ventricular mass / left ventricular end-diastolic volume); CMR, cardiac magnetic resonance imaging; DBP, diastolic blood pressure; EDV, end-diastolic volume; EF, ejection fraction; ESV, end-systolic volume; FD, fractal dimension; HCM, hypertrophic cardiomyopathy; LA, left atrial; LV, left ventricular; LVM, left ventricular mass; LVMI, left ventricular mass index (LVM/BMI); peak diastolic strain rate, PDSR; RA, right atrial; RV, right ventricular; SBP, systolic blood pressure; WT, wall thickness. *Medication for cholesterol, blood pressure, diabetes.

|  | SARC-NEG<br>(n = 288) | SARC-VUS<br>(n = 41) | SARC-P/LP<br>(n = 107) | SARC-NEG vs -P/LP | SARC-NEG vs -VUS | SARC-VUS vs P/LP |
| --- | --- | --- | --- | --- | --- | --- |
| Female (%) | 83 (28.8) | 11 (26.8) | 32 (30.0) |  |  |  |
| Age at scan, y | 60.5 ± 13.4 | 59.9 ± 11.4 | 47.9 ± 14.2 | <0.001 | 0.61 | <0.001 |
| White (n, % of total) | 230 (79.9) | 32 (78.0) | 90 (84.1) |  |  |  |
| BSA, m<sup>2</sup> | 2.0 ± 0.23 | 1.9 ± 0.19 | 2.0 ± 0.4 | | | |
| LVEDV, ml | 136.1 ± 35.8 | 137.3 ± 27.9 | 139.3 ± 37.9 | | | |
| LVESV, ml | 35.9 ± 18.5 | 42.5 ± 18.4 | 38.7 ± 17.9 | 0.06 | 0.03 | 0.253 |
| LVEF, ml | 74.4 ± 8.2 | 69.9 ± 8.3 | 71.8 ± 11.2 | 0.04 | 0.003 | 0.1 |
| LVM, g | 192.7 ± 69.1 | 186.6 ± 48.9 | 172.8 ± 73.2 | 0.005 | 0.85 | 0.03 |
| LVMI, g/m<sup>2</sup> | 98 ± 30.3 | 97.5 ± 23.1 | 88.2 ± 33.6 | <0.001 | 0.6 | 0.005 |
| LV maximum WT, mm | 18.4 ± 4.4 | 18.8 ± 3.5 | 19.3 ± 5.9 | | | |
| Mean apical FD | 1.20 ± 0.05 | 1.22 ± 0.06 | 1.21 ± 0.05 | | | |
| Mean basal FD | 1.20 ± 0.05 | 1.22 ± 0.05 | 1.22 ± 0.05 | <0.001 | 0.106 | 0.009 |
| Mean global FD | 1.2 ± 0.04 | 1.22 ± 0.05 | 1.22 ± 0.04 | 0.001 | 0.008 | 0.948 |
| LV global radial strain, % | 42.0 ± 11.80 | 38.0 ± 10.8 | 41.4 ± 11.39 | | | |
| LV global circumferential strain, % | -19.4 ± 4.03 | -17.4 ± 3.38 | -19.12 ± 4.06 | | | |
| LV radial PDSR | -8.81 ± 2.96 | -8.29 ± 2.86 | -9.41 ± 2.70 | | | |
| LV concentricity g/ml | 1.5 ± 0.5 | 1.4 ± 0.4 | 1.3 ± 0.6 | <0.001 | 0.6 | 0.04 |
| Heart rate, min<sup>-1</sup> | 64.4 ± 24.4 | 62.3 ± 20.5 | 63.5 ± 23.1 | | | |
| Hypertension (n, % of total) | 139 (48.3) | 13 (31.7) | 23 (21.5) | <0.001 | 0.085 | 0.129 |
| On medication* (n, % of total) | 225 (78.1) | 27 (65.9) | 53 (49.5) |  |  |  |
| SBP, mmHg | 122.4 ± 42.8 | 123.5 ± 32.1 | 116.7 ± 41.2 | 0.02 | 0.39 | 0.39 |
| DBP, mmHg | 69.6 ± 24.5 | 72.0 ± 19.7 | 67.8 ± 25.6 | | | |

### Phenotypes associated with rare sarcomeric variants

Unsupervised analysis of patients’ demographic and anthropometric data, clinical characteristics, and cardiac volumes (Supplemental Table 9) was performed using UMAP. Three clusters were identified that were enriched for the following features (Supplemental Fig. 8): i) females with low BSA, moderate hypertrophy and hypertension; ii) males with high BSA, more severe hypertrophy, and hypertension; and iii) younger patients with a family history of HCM, no cardiovascular risk factors, and enrichment for SARC-P/LP variants.

HCM patients with SARC-P/LP variants had lower LV mass (173 ± 73.2 g vs 193 ± 69.1 g, *P* = 0.005) and less concentric remodelling (1.3 ± 0.6 g/ml vs 1.5 ± 0.5 g/ml, *P* < 0.001) than SARC-NEG HCM patients. SARC-P/LP patients also showed increased trabeculation (fractal dimension: 1.22 ± 0.04 vs 1.20 ± 0.04, *P* = 0.001) compared to SARC-NEG patients. SARCNEG HCM patients had a higher prevalence of drug-controlled hypertension than patients with SARC-P/LP variants (48.3% vs 21.5%, *P* < 0.001). Differences in end diastolic and end systolic volumes were not significant.

Three dimensional analysis of cardiac geometry showed that patients with SARC-P/LP variants had a global increase in wall thickness compared to healthy controls predominantly affecting the basal septum (Fig. 2a and b). Within the HCM cohort, SARC-P/LP patients had lower wall thickness across the LV, apart from the basal septum, when compared to those who were SARC-NEG. Although global cavity volumes were similar between genotypes, the three dimensional models showed smaller ventricular cavity size in SARC-NEG individuals in all but the basal septal segments (Fig. 2c, d and e). Taken together these observations suggest that modest global differences in mass and volume between HCM genotypes mask stronger regional variations in geometry and wall thickness.

**Figure 2.**
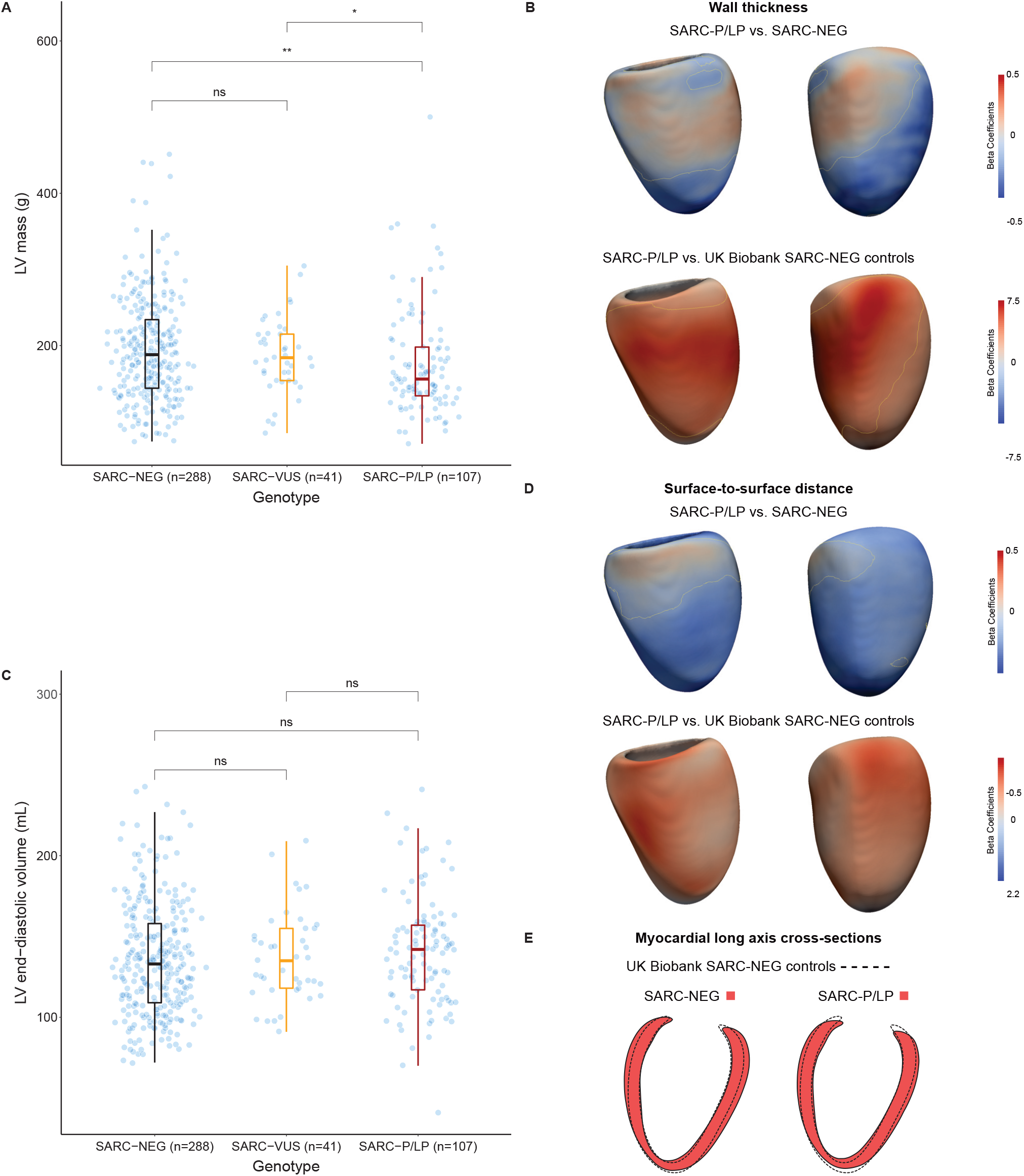
Genotype-phenotype associations in hypertrophic cardiomyopathy. **a**. and **c**. Dot and boxplots of left ventricular (LV) mass and end-diastolic volume in patients with hypertrophic cardiomyopathy stratified by genotype (SARC-NEG, those without variants in genes that may cause or mimic HCM; SARC-P/LP, pathogenic or likely pathogenic sarcomeric variants; SARC-VUS, variants of uncertain significance) **b**. and **d**. 3-dimensional modelling of left ventricular geometry with vertex-wise standardised beta-coefficients projected on the epicardial surface. These show the extent of association between genotype and wall thickness or surface-to-surface distance (comparing regional shape change) for different comparisons, adjusting for the covariates of age, sex and race. Yellow contour lines show significant regions (*P* < 0.05) after multiple testing correction. Left ventricular projections are septal (left) and anterior (right). **e**. Long axis cross sections showing the myocardial outline in red for each genotype compared to control participants in UK Biobank (dashed outline). \**P* ≤ 0.05; \*\**P* ≤ 0.01; ns = not significant.

### Taxonomy of HCM morphology

Individual three dimensional representations of left ventricular morphology were downsampled to form a mesh of 470 vertices points per cardiac phase. DDRtree was used to identify clusters of similar left ventricular geometries and project the data onto a taxonomy where distal branches have more extreme phenotypes and the centre of the tree contains less differentiated phenotypes.^35^ Modelling was performed at both end diastole and end systole to capture structural and functional traits. Significantly associated continuous variables, genotypes, and outcomes are shown for each branch. Internal validation resulted in a good stability of the partitions (Supplemental Tables 1 and 2).

The mapping identified four main phenotypic branches, (Fig. 3, Supplemental Figs. 1 and 4). Variation between these was characterised by comparing average left ventricular morphology in the respective branches to all other patients, and assessing prevalence of genotype status, as well as other imaging and clinical features. Taken together, enriched characteristics in each branch are summarised as: 1) non-sarcomeric mid to apical hypertrophy associated with controlled hypertension and fibrosis; 2) diffuse and basal asymmetric hypertrophy associated with outflow tract obstruction; 3) isolated basal hypertrophy; 4) milder non-obstructive hypertrophy enriched for familial sarcomeric HCM. An additional branch with an undifferentiated pattern of hypertrophy was also identified.

**Figure 3.**
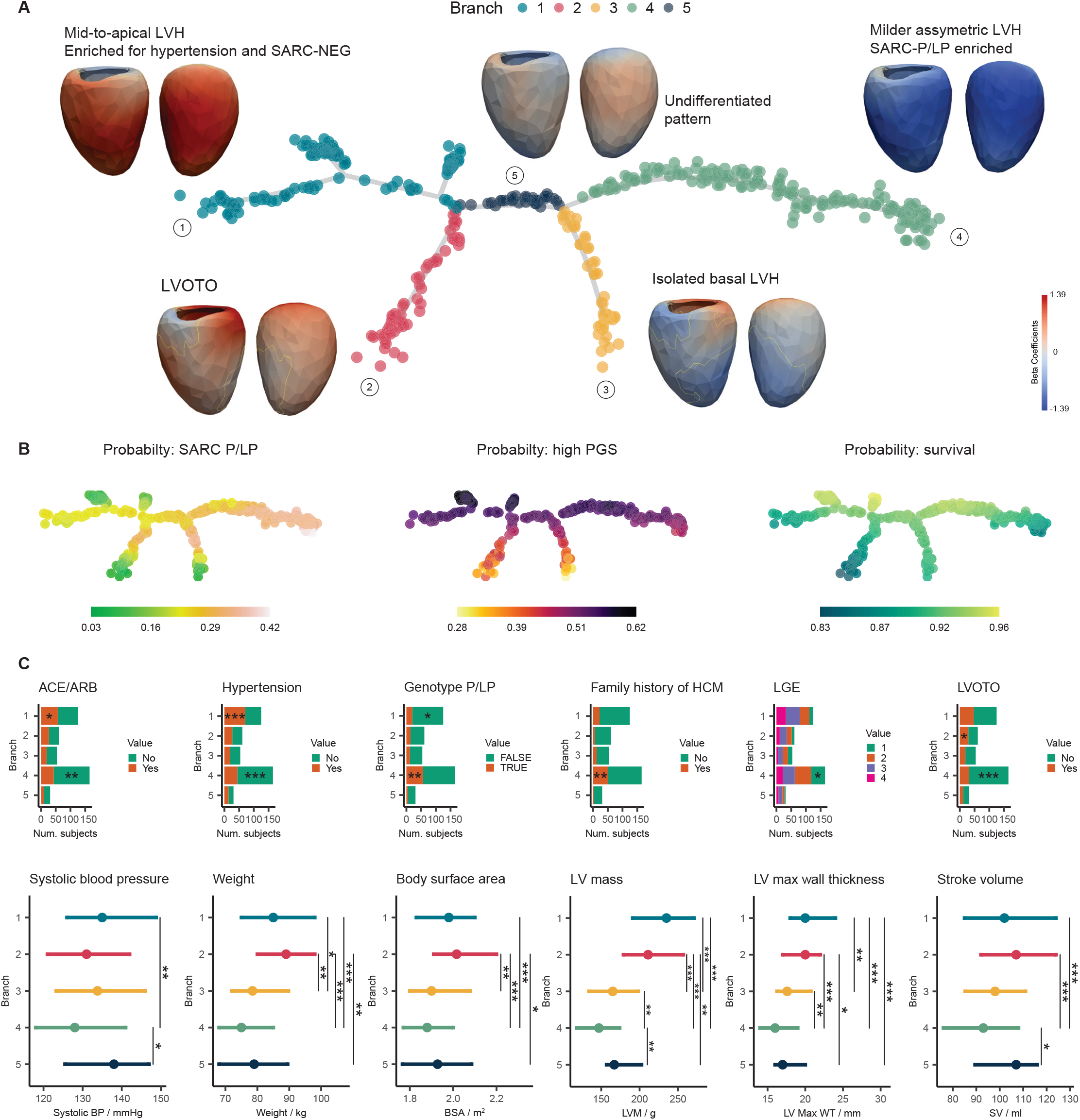
Phenotypic tree of HCM morphology. **a**. Three dimensional models of the left ventricle in patients with HCM were reduced to a two-dimensional tree structure where each point represents one individual. The tree maps undifferentiated states in the centre to more characteristic morphologies in the distal branches while preserving a continuous stratification. For each branch we show a wall thickness shape model at end systole where the colours represent beta coefficients for the comparison between branches. Corresponding features at end diastole are shown in Supplementary Fig. 1. Branch 5 showed an undifferentiated mixed phenotype. **b**. Each participant in the tree labelled by probability of SARC-P/LP genotype, polygenic score (PGS) for HCM, and predicted survival probability at median age. **c**. Continuous and discrete phenotypic variables found to be significantly associated to at least one branch. For late gadolinium enhancement, labels are as follows: 1: None, 2: Minimal, 3: Moderate and 4: Severe. The significance for the enrichment of discrete variables is reported within the bars. ACE, Angiotensin-converting enzyme inhibitors; ARB, Angiotensin receptor blockers; LVOTO, Left ventricular outflow tract obstruction; P/LP, pathogenic or likely pathogenic sarcomeric variants; SV, stroke volume; LGE, late gadolinium enhancement. Only the significant pairs are reported with the symbols: \**P* ≤ 0.05; \*\**P* ≤ 0.01; \*\*\**P* ≤ 0.001; \*\*\*\**P* ≤ 0.0001, n = 436.

We then explored potential causal processes underlying phenotypic heterogeneity and the relationship to outcomes. Phenotypic variation and associated risks can be visualised as a continuous distribution across the whole taxonomic tree, which we show for genotype status, PGS and survival. (Fig. 3B), with end systolic morphology having the greater overall discrimination (Supplemental Table 3 and Supplemental Fig. 5). The tree structure shows how common and rare variants associated with HCM are differentially enriched in the morphological branches and also contribute to a continuous degree of phenotypic expression. For instance, there is enrichment of P/LP, decreased survival, and lower left ventricular mass present on the right side of the tree (branch 4). The median odds ratio for carrying P/LP variants in this branch compared to branch 1 is 2.18 (95% CI: 1.93-2.28, *P* = 0.0001). A higher HCM PGS is associated with SARC-NEG status (*P* = 0.0012) compared to SARC-P/LP. The taxonomy shows a continuous relationship between the predicted probability of high PGS and individual coordinates on the tree as shown in Fig. 3B. Using a logistic regression model with tree coordinates as independent variables we observed an association between the vertical axis of the tree and PGS (*P* = 0.0025) showing lower PGS in more differentiated isolated basal LVH and LVOTO-enriched phenotypes.

### External validation

New patients with imaging can be mapped to coordinates in the tree structure to visualise individual risk and morphological differentiation. We mapped an independent external cohort of HCM patients with CMR imaging and assessed similarity of phenotypic patterns. Predictive random forest models for the 2 tree coordinates were tested on the development cohort with 10-fold cross-validation, repeated 3 times. Both models had good performances (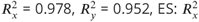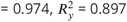, for x and y coordinates, respectively). These models were used to predict the tree coordinates of the external HCM cases from their wall thickness values adjusted for sex and age at scan.

Visual inspection of the predicted tree coordinates showed no outliers, with the points falling close to the tree structure (Supplemental Fig. 6). Branch labels were assigned from the nearest neighbor in the development cohort (Supplemental Table 4). This preserved the faithfulness of the mapping, as confirmed by the estimated “trustworthiness”^36^ (see Supplementary material) (*M*_*1*_ - ED range: 0.83-0.87, ES range: 0.86-0.88). No significant difference was found in the distribution of correlations within two cohorts supporting consistency between their projections on the tree (Supplemental Fig. 7)

### Clinical outcomes

All-cause mortality was available for all 436 HCM cases (Supplementary Fig. 9) and was found to be 17.4% across the entire cohort. By the end of follow up, 14 (13.1%) of the SARC-P/LP, 7 (17.1%) of the SARC-VUS and 55 (19.1%) of the SARC-NEG patients had died. The SARC-P/LP patients were younger at recruitment (median age 49 years, IQR: 38-61); *P* < 0.001) than both SARC-VUS (median age 61 years, IQR 52-67) and SARC-NEG patients (median age 62 years, IQR: 53-71). In an unadjusted Cox model, SARC-P/LP patients had increased risk of death (n = 395, HR: 2.66; 95% CI: 1.42-4.96; *P* = 0.002; Kaplan-Meier plot in Supplementary Fig. 9) relative to SARC-NEG participants and shorter lifespan (median age at death 67 years, IQR: 57-70 vs 76 years, IQR: 68-84; *P* = 0.0003). This relationship was also independent of genetic sex and ancestry in a multivariable analysis (Supplemental Tables 7 and 8).

## Discussion

Hypertrophic cardiomyopathy is characterised by phenotypic heterogeneity, which presents challenges for developing personalised profiles of dynamic disease status to guide patient management. Conventional morphological descriptions of hypertrophic remodelling have relied on two dimensional appearances with a diverse range of subjective shape classifications proposed with varying genotype enrichment.^37,38^ Here we report a systematic study of three dimensional genotypephenotype associations in sarcomeric and non-sarcomeric HCM, and identify natural groupings of morphologically similar patients while preserving the continuous distribution of phenotypic severity, clinical risk and enrichment for HCM-associated rare variants and common variant modifiers.

The classification of HCM morphology has relied on recognising regional distributions and shape features of left ventricular hypertrophy, initially with echocardiography and more recently using CMR, with between 6 to 12 different patterns described.^39–41^ Broadly, these comprise basal (sigmoid), mid-ventricular (reverse septal curve), apical, and diffuse patterns, as well as mixed phenotypes. Within these heterogeneous endophenotypes, those with SARC-P/LP variants are more likely to have reverse septal curve morphology with fibrosis, and those who are SARC-NEG more likely to have isolated basal septal hypertrophy with obstruction but less fibrosis.^7^ While such approaches either associate imaging phenotypes that share common features or stratify by genotype status, they do not objectively classify the spectrum of phenotypic expression that is characteristic of HCM. Advances in image analysis have allowed detailed 3D models of ventricular shape and motion to be extracted from routine CMR that possess spatial consistency within and between cohorts, allowing precise aggregation of phenotypic data in diverse populations.^5^ This enables regional genotype-phenotype associations to be made by variant status and allows data-driven approaches for unsupervised learning of complex traits.

In our study, SARC-NEG HCM patients had increased LV mass compared to SARC-P/LP patients but this masked differences in the patterns of hypertrophy between genotypes. Three dimensional genotype-phenotype mapping showed that SARC-P/LP patients had lower wall thickness across most of the left ventricle, apart from the basal septum, when compared to HCM patients who were SARC-NEG. Relative to healthy controls there was diffuse hypertrophy that was most marked in the basal septum. Cross sections of the left ventricle show a generally smaller end diastolic cavity in SARC-P/LP patients compared to controls, especially in the long axis direction, while SARC-NEG HCM patients have a similar cavity size to controls but show more diffuse hypertrophy. Differences in global volumes and mass between genotypes are therefore the net effect of opposing regional changes in the distribution and degree of hypertrophy.

To reconstruct trajectories of how HCM phenotypes transition towards differentiated states we applied a machine learning approach, using reversed graph embedding, to learn branch points that define significant divergences in 3D morphology that requires no *a priori* information about the genes or environment that modify disease biology.^42^ The resulting tree structure provides a two-dimensional representation of complex phenotypic variation that can be generalised to diverse datasets,^35^ and addresses potential sources of bias and subjectivity in human visual assessment of HCM.^43^ The tree maps disease trajectories from undifferentiated states in the centre to more characteristic morphologies in the distal branches. This provides a visual representation of how phenotypic variation translates to variation in genetic enrichment and survival. We also demonstrate how new patients can be mapped to a position in the tree, facilitating individualised patient stratification.

The interaction between hypertension and HCM presents significant diagnostic challenges, including when it should be considered a co-morbid condition rather than the underlying cause of hypertrophy.^10^ The phenotypic branch with the most severe degree of hypertrophy (1) was enriched for controlled hypertension and comprised predominately SARC-NEG patients. The pattern of hypertrophy predominantly affects the mid-ventricle to apex in this branch and may include the phenotypic spectrum of “apical” HCM.^44^ Non-sarcomeric HCM is partly an exaggerated response to diastolic hypertension in genetically susceptible individuals and this phenotype shares features of hypertensive patients without HCM.^3,45^ Late enhancement burden was highest in this group, suggesting that enrichment for replacement fibrosis is associated with more severe manifestations of HCM despite a lower prevalence of pathogenic variants.^46^ Recognition of these patients’ phenotype at the point of diagnosis could help to identify non-sarcomeric HCM cases associated with modifiable risk factors. The other common phenotypic branch (4) had milder hypertrophy, a low prevalence of hypertension and the most enrichment for P/LP variants. In common with previous registry studies, we also found that P/LP variants to be predictors of mortality,^47^ and this non-obstructive HCM phenotype enriched for familial disease and sarcomeric variants, had the poorest outcome with risk increasing for patients expressing the most differentiated phenotype in the tree structure. Early prediction of molecular sub-types of HCM could be of value when considering emerging therapies that act upstream of the underlying genetic cause.^48^

Left ventricular outflow tract obstruction (LVOTO) is independently associated with adverse HCM-related outcomes,^49^ may benefit from surgical intervention, and is amenable to treatment with myosin inhibitors.^50^ It is associated with septal hypertrophy and ventricular remodelling,^51^ although it may also occur in milder cases of HCM.^52^ We found that the phenotype in branch (2) with pronounced mid to basal asymmetric hypertrophy in a diffusely hypertrophic left ventricle was strongly enriched for stress-confirmed LVOTO and hyperdynamic function. As this phenotype becomes more differentiated from an average HCM morphology the survival probability decreases. The last main group (3) in the taxonomy is associated with isolated hypertrophy of the basal septum with relatively mild left ventricular hypertrophy elsewhere. A similar benign phenotype has been recognised as part of normal aging whose appearances may overlap with HCM.^53^ We also found this phenotype to be associated with better survival and a low probability of P/LP variants.

Our approach differs from previous studies describing phenotypic patterns in HCMs as we adjust for age and sex, which affect both disease physiology and outcomes, and so the classification is independent of these risk factors.^54^ While our approach also aims to identify groups of similar morphology it does this using a data-driven approach without human subjectivity and maintains a continuum of disease expression and risk factors across the taxonomic tree. We also identify potential causal modifiers of disease expression with common variants contributing to the differentiation and severity of phenotypes in addition to their reported role as risk markers for survival and adverse events in HCM.^27^ The current model is not intended to guide treatment at present, but offers an approach to understand a complex genotype-phenotype architecture in a comprehensible model for comparison of heterogeneous disease states.^35^ We also show how unseen external data can be readily mapped to coordinates in the tree to explore personal dynamic risk profiles. Multiparametric imaging-based models offer more accurate prediction of composite endpoints in HCM compared to conventional risk factors,^55^ and machine learning models using conventional imaging parameters have better discrimination of HCM genotype than clinical scores.^56^ The full value of these concepts could be realised across diverse disease states, including integration of imaging with multi-omic profiles, to provide a comprehensive phenotypic and molecular classification of cardiomyopathy. This study has limitations. The HCM biobank recruited consecutive patients with a clinical diagnosis from a cardiomyopathy clinic or undergoing CMR, and so may be less enriched for familial, symptomatic or more severe phenotypes compared to other cohorts. This reflects the spectrum of disease seen in clinical practice but may account for the relatively low event rate observed. Most of the development cohort was European, although we found that the mapping could generalise well to other ancestries. We used CMR as this is regarded as the gold standard for phenotyping, but it is less widely available than echocardiography.

In summary, we provide a data-driven taxonomy for understanding dynamic phenotypic diversity in HCM that reflects a continuum of disease, genetic risk and outcomes. A systematic representation HCM diversity onto which new patients can be mapped has the potential to enable more individualised assessment, stratification and treatment strategies.

## Data Availability

The code used in the study is available on GitHub. Population data is available from UK
Biobank.

https://github.com/ImperialCollegeLondon/HCM-taxonomy

## Author contributions

L.C. and A. de M. performed data curation and analysis, and drafted the manuscript; P.I., A.C. and P-R.S. performed analysis and visualisation of the data; K.A.McG., R.J.B. and P.T. performed the genetic analysis, S.L., M.J. and W.B. performed the image analysis; M.S. and S.L.Z. performed formal analysis of the data; C.E.R., A.J.P, A.P. and B.P.H. collected clinical data in the U.K.; C.J.P. and C.W.L.C. collected clinical and genetic data in Singapore; D.J.P., S.A.C., S.K.P., S.J.J, R.T, C.R.B., and H.W. provided clinical or genetic resources for the study; J.S.W. supervised the genetic analyses; D.P.O’R. conceived the project, provided supervision and funding. All authors contributed to the article and approved the submitted version.

## Data availability

The code used in the study is available on GitHub (https://github.com/ImperialCollegeLondon/HCM-taxonomy; doi:10.5281/zenodo.7639435). Population data is available from UK Biobank.

## Acknowledgments

We thank Dr Tim Dawes and Dr Carlo Biffi (Imperial College London) for contributing to the initial development of the three dimensional cardiac modelling. We also acknowledge Dr Jinming Duan (University of Birmingham) in the initial development of the segmentation pipeline.

## References

1. Maron BJ, Desai MY, Nishimura RA, Spirito P, Rakowski H, Towbin JA, Rowin EJ, Maron MS & Sherrid MV. Diagnosis and Evaluation of Hypertrophic Cardiomyopathy: JACC State-of-the-Art Review. J Am Coll Cardiol 79, 372–389. doi:10.1016/j.jacc.2021.12.002 (2022).

2. Tadros R, Francis C, Xu X, Vermeer AMC, Harper AR, Huurman R, Kelu Bisabu K, Walsh R, Hoorntje ET, Te Rijdt WP, Buchan RJ, van Velzen HG, van Slegtenhorst MA, Vermeulen JM, Offerhaus JA, Bai W, de Marvao A, Lahrouchi N, Beekman L, Karper JC, Veldink JH, Kayvanpour E, Pantazis A, Baksi AJ, Whiffin N, Mazzarotto F, Sloane G, Suzuki H, Schneider-Luftman D, Elliott P, Richard P, Ader F, Villard E, Lichtner P, Meitinger T, Tanck MWT, van Tintelen JP, Thain A, McCarty D, Hegele RA, Roberts JD, Amyot J, Dubé MP, Cadrin-Tourigny J, Giraldeau G, L’Allier PL, Garceau P, Tardif JC, Boekholdt SM, Lumbers RT, Asselbergs FW, Barton PJR, Cook SA, Prasad SK, O’Regan DP, van der Velden J, Verweij KJH, Talajic M, Lettre G, Pinto YM, Meder B, Charron P, de Boer RA, Christiaans I, Michels M, Wilde AAM, Watkins H, Matthews PM, Ware JS & Bezzina CR. Shared genetic pathways contribute to risk of hypertrophic and dilated cardiomyopathies with opposite directions of effect. Nat Genet 53, 128–134. doi:10.1038/s41588-020-00762-2 (2021).

3. Harper AR, Goel A, Grace C, Thomson KL, Petersen SE, Xu X, Waring A, Ormondroyd E, Kramer CM, Ho CY, Neubauer S, Investigators H, Tadros R, Ware JS, Bezzina CR, Farrall M & Watkins H. Common genetic variants and modifiable risk factors underpin hypertrophic cardiomyopathy susceptibility and expressivity. Nat Genet 53, 135–142. doi:10.1038/s41588-020-00764-0 (2021).

4. Butters A, Lakdawala NK & Ingles J. Sex Differences in Hypertrophic Cardiomyopathy: Interaction With Genetics and Environment. Curr Heart Fail Rep 18, 264–273. doi:10.1007/s11897-021-00526-x (2021).

5. De Marvao A, McGurk KA, Zheng SL, Thanaj M, Bai W, Duan J, Biffi C, Mazzarotto F, Statton B, Dawes TJW, Savioli N, Halliday BP, Xu X, Buchan RJ, Baksi AJ, Quinlan M, Tokarczuk P, Tayal U, Francis C, Whiffin N, Theotokis PI, Zhang X, Jang M, Berry A, Pantazis A, Barton PJR, Rueckert D, Prasad SK, Walsh R, Ho CY, Cook SA, Ware JS & O’Regan DP. Phenotypic Expression and Outcomes in Individuals With Rare Genetic Variants of Hypertrophic Cardiomyopathy. J Am Coll Cardiol 78, 1097–1110. doi:10.1016/j.jacc.2021.07.017 (2021).

6. Raman B, Ariga R, Spartera M, Sivalokanathan S, Chan K, Dass S, Petersen SE, Daniels MJ, Francis J, Smillie R, Lewandowski AJ, Ohuma EO, Rodgers C, Kramer CM, Mahmod M, Watkins H & Neubauer S. Progression of myocardial fibrosis in hypertrophic cardiomyopathy: mechanisms and clinical implications. Eur Heart J Cardiovasc Imaging 20, 157–167. doi:10.1093/ehjci/jey135 (2019).

7. Neubauer S, Kolm P, Ho CY, Kwong RY, Desai MY, Dolman SF, Appelbaum E, Desvigne-Nickens P, DiMarco JP, Friedrich MG, Geller N, Harper AR, Jarolim P, Jerosch-Herold M, Kim DY, Maron MS, Schulz-Menger J, Piechnik SK, Thomson K, Zhang C, Watkins H, Weintraub WS & Kramer CM. Distinct Subgroups in Hypertrophic Cardiomyopathy in the NHLBI HCM Registry. J Am Coll Cardiol 74, 2333–2345. doi:10.1016/j.jacc.2019.08.1057 (2019).

8. Piras P, Torromeo C, Evangelista A, Esposito G, Nardinocchi P, Teresi L, Madeo A, Re F, Chialastri C, Schiariti M, Varano V & Puddu PE. Non-invasive prediction of genotype positive-phenotype negative in hypertrophic cardiomyopathy by 3D modern shape analysis. Exp Physiol 104, 1688–1700. doi:10.1113/ep087551 (2019).

9. Maron BJ, Maron MS, Maron BA & Loscalzo J. Moving Beyond the Sarcomere to Explain Heterogeneity in Hypertrophic Cardiomyopathy: JACC Review Topic of the Week. J Am Coll Cardiol 73, 1978–1986. doi:10.1016/j.jacc.2019.01.061 (2019).

10. Ommen SR, Mital S, Burke MA, Day SM, Deswal A, Elliott P, Evanovich LL, Hung J, Joglar JA, Kantor P, Kimmelstiel C, Kittleson M, Link MS, Maron MS, Martinez MW, Miyake CY, Schaff HV, Semsarian C & Sorajja P. 2020 AHA/ACC Guideline for the Diagnosis and Treatment of Patients With Hypertrophic Cardiomyopathy: A Report of the American College of Cardiology/American Heart Association Joint Committee on Clinical Practice Guidelines. Circulation 142, e558–e631. doi:10.1161/cir.0000000000000937 (2020).

11. Gersh BJ, Maron BJ, Bonow RO, Dearani JA, Fifer MA, Link MS, Naidu SS, Nishimura RA, Ommen SR, Rakowski H, Seidman CE, Towbin JA, Udelson JE & Yancy CW. 2011 ACCF/AHA guideline for the diagnosis and treatment of hypertrophic cardiomyopathy: executive summary: a report of the American College of Cardiology Foundation/American Heart Association Task Force on Practice Guidelines. Circulation 124, 2761–96. doi:10.1161/CIR.0b013e318223e230 (2011).

12. Bycroft C, Freeman C, Petkova D, Band G, Elliott LT, Sharp K, Motyer A, Vukcevic D, Delaneau O, O’Connell J, Cortes A, Welsh S, Young A, Effingham M, McVean G, Leslie S, Allen N, Donnelly P & Marchini J. The UK Biobank resource with deep phenotyping and genomic data. Nature 562, 203–209. doi:10.1038/s41586-018-0579-z (2018).

13. Petersen SE, Matthews PM, Francis JM, Robson MD, Zemrak F, Boubertakh R, Young AA, Hudson S, Weale P, Garratt S, Collins R, Piechnik S & Neubauer S. UK Biobank’s cardiovascular magnetic resonance protocol. J Cardiovasc Magn Reson 18, 8. doi:10.1186/s12968-016-0227-4 (2016).

14. Bai W, Sinclair M, Tarroni G, Oktay O, Rajchl M, Vaillant G, Lee AM, Aung N, Lukaschuk E, Sanghvi MM, Zemrak F, Fung K, Paiva JM, Carapella V, Kim YJ, Suzuki H, Kainz B, Matthews PM, Petersen SE, Piechnik SK, Neubauer S, Glocker B & Rueckert D. Automated cardiovascular magnetic resonance image analysis with fully convolutional networks. J Cardiovasc Magn Reson 20, 65. doi:10.1186/s12968-018-0471-x (2018).

15. Duan J, Bello G, Schlemper J, Bai W, Dawes TJW, Biffi C, de Marvao A, Doumoud G, O’Regan DP & Rueckert D. Automatic 3D Bi-Ventricular Segmentation of Cardiac Images by a Shape-Refined Multi-Task Deep Learning Approach. IEEE Trans Med Imaging 38, 2151–2164. doi:10.1109/tmi.2019.2894322 (2019).

16. Schulz-Menger J, Bluemke DA, Bremerich J, Flamm SD, Fogel MA, Friedrich MG, Kim RJ, von Knobelsdorff-Brenkenhoff F, Kramer CM, Pennell DJ, Plein S & Nagel E. Standardized image interpretation and post-processing in cardiovascular magnetic resonance - 2020 update : Society for Cardiovascular Magnetic Resonance (SCMR): Board of Trustees Task Force on Standardized Post-Processing. J Cardiovasc Magn Reson 22, 19. doi:10.1186/s12968-020-00610-6 (2020).

17. Puyol-Antón E, Ruijsink B, Bai W, Langet H, De Craene M, Schnabel JA, Piro P, King AP & Sinclair M. Fully automated myocardial strain estimation from cine MRI using convolutional neural networks in 2018 IEEE 15th International Symposium on Biomedical Imaging (ISBI 2018) (2018), 1139–1143. doi:10.1109/ISBI.2018.8363772.

18. Meyer HV, Dawes TJW, Serrani M, Bai W, Tokarczuk P, Cai J, de Marvao A, Henry A, Lumbers RT, Gierten J, Thumberger T, Wittbrodt J, Ware JS, Rueckert D, Matthews PM, Prasad SK, Costantino ML, Cook SA, Birney E & O’Regan DP. Genetic and functional insights into the fractal structure of the heart. Nature 584, 589–594. doi:10.1038/s41586-020-2635-8 (2020).

19. Walsh R, Buchan R, Wilk A, John S, Felkin LE, Thomson KL, Chiaw TH, Loong CCW, Pua CJ, Raphael C, Prasad S, Barton PJ, Funke B, Watkins H, Ware JS & Cook SA. Defining the genetic architecture of hypertrophic cardiomyopathy: reevaluating the role of non-sarcomeric genes. Eur Heart J 38, 3461–3468. doi:10.1093/eurheartj/ehw603 (2017).

20. Pua CJ, Tham N, Chin CWL, Walsh R, Khor CC, Toepfer CN, Repetti GG, Garfinkel AC, Ewoldt JF, Cloonan P, Chen CS, Lim SQ, Cai J, Loo LY, Kong SC, Chiang CWK, Whiffin N, de Marvao A, Lio PM, Hii AA, Yang CX, L. TT, Bylstra Y, Lim WK, Teo JX, Padilha K, Silva GV, Pan B, Govind R, Buchan RJ, Barton PJR, Tan P, Foo R, Yip JWL, Wong RCC, Chan WX, Pereira AC, Tang HC, Jamuar SS, Ware JS, Seidman JG, Seidman CE & Cook SA. Genetic Studies of Hypertrophic Cardiomyopathy in Singaporeans Identify Variants in TNNI3 and TNNT2 That Are Common in Chinese Patients. Circ Genom Precis Med 13, 424–434. doi:10.1161/circgen.119.002823 (2020).

21. Whiffin N, Minikel E, Walsh R, O’Donnell-Luria AH, Karczewski K, Ing AY, Barton PJR, Funke B, Cook SA, MacArthur D & Ware JS. Using high-resolution variant frequencies to empower clinical genome interpretation. Genet Med 19, 1151–1158. doi:10.1038/gim.2017.26 (2017).

22. Whiffin N, Walsh R, Govind R, Edwards M, Ahmad M, Zhang X, Tayal U, Buchan R, Midwinter W, Wilk AE, Najgebauer H, Francis C, Wilkinson S, Monk T, Brett L, O’Regan DP, Prasad SK, Morris-Rosendahl DJ, Barton PJR, Edwards E, Ware JS & Cook SA. CardioClassifier: disease- and gene-specific computational decision support for clinical genome interpretation. Genet Med 20, 1246–1254. doi:10.1038/gim.2017.258 (2018).

23. Walsh R, Mazzarotto F, Whiffin N, Buchan R, Midwinter W, Wilk A, Li N, Felkin L, Ingold N, Govind R, Ahmad M, Mazaika E, Allouba M, Zhang X, de Marvao A, Day SM, Ashley E, Colan SD, Michels M, Pereira AC, Jacoby D, Ho CY, Thomson KL, Watkins H, Barton PJR, Olivotto I, Cook SA & Ware JS. Quantitative approaches to variant classification increase the yield and precision of genetic testing in Mendelian diseases: the case of hypertrophic cardiomyopathy. Genome Med 11, 5. doi:10.1186/s13073-019-0616-z (2019).

24. Richards S, Aziz N, Bale S, Bick D, Das S, Gastier-Foster J, Grody WW, Hegde M, Lyon E, Spector E, Voelkerding K & Rehm HL. Standards and guidelines for the interpretation of sequence variants: a joint consensus recommendation of the American College of Medical Genetics and Genomics and the Association for Molecular Pathology. Genet Med 17, 405–24. doi:10.1038/gim.2015.30 (2015).

25. Karczewski KJ, Francioli LC, Tiao G, Cummings BB, Alföldi J, Wang Q, Collins RL, Laricchia KM, Ganna A, Birnbaum DP, Gauthier LD, Brand H, Solomonson M, Watts NA, Rhodes D, Singer-Berk M, England EM, Seaby EG, Kosmicki JA, Walters RK, Tashman K, Farjoun Y, Banks E, Poterba T, Wang A, Seed C, Whiffin N, Chong JX, Samocha KE, Pierce-Hoffman E, Zappala Z, O’Donnell-Luria AH, Minikel EV, Weisburd B, Lek M, Ware JS, Vittal C, Armean IM, Bergelson L, Cibulskis K, Connolly KM, Covarrubias M, Donnelly S, Ferriera S, Gabriel S, Gentry J, Gupta N, Jeandet T, Kaplan D, Llanwarne C, Munshi R, Novod S, Petrillo N, Roazen D, Ruano-Rubio V, Saltzman A, Schleicher M, Soto J, Tibbetts K, Tolonen C, Wade G, Talkowski ME, Neale BM, Daly MJ & MacArthur DG. The mutational constraint spectrum quantified from variation in 141,456 humans. Nature 581, 434–443. doi:10.1038/s41586-020-2308-7 (2020).

26. Ingles J, Goldstein J, Thaxton C, Caleshu C, Corty EW, Crowley SB, Dougherty K, Harrison SM, McGlaughon J, Milko LV, Morales A, Seifert BA, Strande N, Thomson K, Peter van Tintelen J, Wallace K, Walsh R, Wells Q, Whiffin N, Witkowski L, Semsarian C, Ware JS, Hershberger RE & Funke B. Evaluating the Clinical Validity of Hypertrophic Cardiomyopathy Genes. Circ Genom Precis Med 12, e002460–e002460. doi:10.1161/CIRCGEN.119.002460 (2019).

27. Zheng SL, Jurgens SJ, McGurk KA, Xu X, Grace C, Theotokis PI, Buchan R, Francis C, de Marvao A, Curran L, et al. Evaluation of polygenic score for hypertrophic cardiomyopathy in the general population and across clinical settings. medRxiv. doi:10.1101/2023.03.14.23286621 (2023).

28. Sainburg T, McInnes L & Gentner TQ. Parametric UMAP Embeddings for Representation and Semisupervised Learning. Neural Comput 33, 2881–2907. doi:10.1162/neco_a_01434 (2021).

29. Biffi C, de Marvao A, Attard MI, Dawes TJW, Whiffin N, Bai W, Shi W, Francis C, Meyer H, Buchan R, Cook SA, Rueckert D & O’Regan DP. Three-dimensional cardiovascular imaging-genetics: a mass univariate framework. Bioinformatics 34, 97–103. doi:10.1093/bioinformatics/btx552 (2018).

30. Jarvis R & Patrick E. Clustering Using a Similarity Measure Based on Shared Near Neighbors. IEEE Trans Comput C-22, 1025–1034. doi:10.1109/T-C.1973.223640 (1973).

31. Waltman L & van Eck NJ. A smart local moving algorithm for large-scale modularity-based community detection. Eur Phys J B 86. doi:10.1140/epjb/e2013-40829-0 (2013).

32. Hao Y, Hao S, Andersen-Nissen E, Mauck W. M.3, Zheng S, Butler A, Lee MJ, Wilk AJ, Darby C, Zager M, Hoffman P, Stoeckius M, Papalexi E, Mimitou EP, Jain J, Srivastava A, Stuart T, Fleming LM, Yeung B, Rogers AJ, McElrath JM, Blish CA, Gottardo R, Smibert P & Satija R. Integrated analysis of multimodal single-cell data. Cell 184, 3573–3587.e29. doi:10.1016/j.cell.2021.04.048 (2021).

33. Hennig C. fpc: Flexible Procedures for Clustering R package version 2.2-9 (2020). https://CRAN.R-project.org/package=fpc.

34. Mao Q, Wang L, Goodison S & Sun Y. Dimensionality Reduction Via Graph Structure Learning in Proceedings of the 21th ACM SIGKDD International Conference on Knowledge Discovery and Data Mining (2015), 765–774. doi:10.1145/2783258.2783309.

35. Nair ATN, Wesolowska-Andersen A, Brorsson C, Rajendrakumar AL, Hapca S, Gan S, Dawed AY, Donnelly LA, McCrimmon R, Doney AS, et al. Heterogeneity in phenotype, disease progression and drug response in type 2 diabetes. Nat Med 28, 982–988. doi:10.1038/s41591-022-01790-7 (2022).

36. Van der Maaten L. Learning a Parametric Embedding by Preserving Local Structure in Proceedings of the Twelfth International Conference on Artificial Intelligence and Statistics (eds van Dyk D & Welling M) 5 (PMLR, Hilton Clearwater Beach Resort, Clearwater Beach, Florida USA, 2009), 384–391.

37. Olivotto I, Cecchi F, Poggesi C & Yacoub MH. Patterns of disease progression in hypertrophic cardiomyopathy: an individualized approach to clinical staging. Circ Heart Fail 5, 535–46. doi:10.1161/circheartfailure.112.967026 (2012).

38. Binder J, Ommen SR, Gersh BJ, Van Driest SL, Tajik AJ, Nishimura RA & Ackerman MJ. Echocardiography-guided genetic testing in hypertrophic cardiomyopathy: septal morphological features predict the presence of myofilament mutations. Mayo Clin Proc 81, 459–67. doi:10.4065/81.4.459 (2006).

39. Klues HG, Schiffers A & Maron BJ. Phenotypic spectrum and patterns of left ventricular hypertrophy in hypertrophic cardiomyopathy: morphologic observations and significance as assessed by two-dimensional echocardiography in 600 patients. J Am Coll Cardiol 26, 1699–708. doi:10.1016/0735-1097(95)00390-8 (1995).

40. Maron MS. Clinical utility of cardiovascular magnetic resonance in hypertrophic cardiomyopathy. J Cardiovasc Magn Reson 14, 13. doi:10.1186/1532-429x-14-13 (2012).

41. Maron MS, Maron BJ, Harrigan C, Buros J, Gibson CM, Olivotto I, Biller L, Lesser JR, Udelson JE, Manning WJ & Appelbaum E. Hypertrophic cardiomyopathy phenotype revisited after 50 years with cardiovascular magnetic resonance. J Am Coll Cardiol 54, 220–8. doi:10.1016/j.jacc.2009.05.006 (2009).

42. Qiu X, Mao Q, Tang Y, Wang L, Chawla R, Pliner HA & Trapnell C. Reversed graph embedding resolves complex singlecell trajectories. Nat Methods 14, 979–982. doi:10.1038/nmeth.4402 (2017).

43. Augusto JB, Davies RH, Bhuva AN, Knott KD, Seraphim A, Alfarih M, Lau C, Hughes RK, Lopes LR, Shiwani H, Treibel TA, Gerber BL, Hamilton-Craig C, Ntusi NAB, Pontone G, Desai MY, Greenwood JP, Swoboda PP, Captur G, Cavalcante J, Bucciarelli-Ducci C, Petersen SE, Schelbert E, Manisty C & Moon JC. Diagnosis and risk stratification in hypertrophic cardiomyopathy using machine learning wall thickness measurement: a comparison with human test-retest performance. Lancet Digit Health 3, e20–e28. doi:10.1016/s2589-7500(20)30267-3 (2021).

44. Hughes RK, Knott KD, Malcolmson J, Augusto JB, Mohiddin SA, Kellman P, Moon JC & Captur G. Apical Hypertrophic Cardiomyopathy: The Variant Less Known. J Am Heart Assoc 9, e015294. doi:10.1161/JAHA.119.015294 (2020).

45. De Marvao A, Dawes TJ, Shi W, Durighel G, Rueckert D, Cook SA & O’Regan DP. Precursors of Hypertensive Heart Phenotype Develop in Healthy Adults: A High-Resolution 3D MRI Study. JACC Cardiovasc Imaging 8, 1260–9. doi:10.1016/j.jcmg.2015.08.007 (2015).

46. Liu J, Zhao S, Yu S, Wu G, Wang D, Liu L, Song J, Zhu Y, Kang L, Wang J & Song L. Patterns of Replacement Fibrosis in Hypertrophic Cardiomyopathy. Radiology 302, 298–306. doi:10.1148/radiol.2021210914 (2022).

47. Ho CY, Day SM, Ashley EA, Michels M, Pereira AC, Jacoby D, Cirino AL, Fox JC, Lakdawala NK, Ware JS, Caleshu CA, Helms AS, Colan SD, Girolami F, Cecchi F, Seidman CE, Sajeev G, Signorovitch J, Green EM & Olivotto I. Genotype and Lifetime Burden of Disease in Hypertrophic Cardiomyopathy: Insights from the Sarcomeric Human Cardiomyopathy Registry (SHaRe). Circulation 138, 1387–1398. doi:10.1161/circulationaha.117.033200 (2018).

48. Helms AS, Thompson AD & Day SM. Translation of New and Emerging Therapies for Genetic Cardiomyopathies. JACC Basic Transl Sci 7, 70–83. doi:10.1016/j.jacbts.2021.07.012 (2022).

49. Maron MS, Olivotto I, Betocchi S, Casey SA, Lesser JR, Losi MA, Cecchi F & Maron BJ. Effect of left ventricular outflow tract obstruction on clinical outcome in hypertrophic cardiomyopathy. N Engl J Med 348, 295–303. doi:10.1056/NEJMoa021332 (2003).

50. Spertus JA, Fine JT, Elliott P, Ho CY, Olivotto I, Saberi S, Li W, Dolan C, Reaney M, Sehnert AJ & Jacoby D. Mavacamten for treatment of symptomatic obstructive hypertrophic cardiomyopathy (EXPLORER-HCM): health status analysis of a randomised, double-blind, placebo-controlled, phase 3 trial. Lancet 397, 2467–2475. doi:10.1016/S0140-6736(21)00763-7 (2021).

51. Hermida U, Stojanovski D, Raman B, Ariga R, Young AA, Carapella V, Carr-White G, Lukaschuk E, Piechnik SK, Kramer CM, Desai MY, Weintraub WS, Neubauer S, Watkins H & Lamata P. Left ventricular anatomy in obstructive hypertrophic cardiomyopathy: beyond basal septal hypertrophy. Eur Heart J Cardiovasc Imaging. doi:10.1093/ehjci/jeac233 (2022).

52. Patel P, Dhillon A, Popovic ZB, Smedira NG, Rizzo J, Thamilarasan M, Agler D, Lytle BW, Lever HM & Desai MY. Left Ventricular Out?ow Tract Obstruction in Hypertrophic Cardiomyopathy Patients Without Severe Septal Hypertrophy: Implications of Mitral Valve and Papillary Muscle Abnormalities Assessed Using Cardiac Magnetic Resonance and Echocardiography. Circ Cardiovasc Imaging 8, e003132. doi:10.1161/CIRCIMAGING.115.003132 (2015).

53. Diaz T, Pencina MJ, Benjamin EJ, Aragam J, Fuller DL, Pencina KM, Levy D & Vasan RS. Prevalence, clinical correlates, and prognosis of discrete upper septal thickening on echocardiography: the Framingham Heart Study. Echocardiography 26, 247–53. doi:10.1111/j.1540-8175.2008.00806.x (2009).

54. Lee HJ, Kim HK, Lee SC, Ommen SR, Kim J, Park JB, Choi YJ, Lee SP, Chang SA & Kim YJ. Age-related sex differences in the outcomes of patients with hypertrophic cardiomyopathy. PLoS One 17, e0264580. doi:10.1371/journal.pone.0264580 (2022).

55. Georgiopoulos G, Figliozzi S, Pateras K, Nicoli F, Bampatsias D, Beltrami M, Finocchiaro G, Chiribiri A, Masci PG & Olivotto I. Comparison of demographic, clinical, biochemical, and imaging findings in hypertrophic cardiomyopathy prognosis: a network meta-analysis. JACC Heart Fail 11, 30–41. doi:10.1016/j.jchf.2022.08.022 (2022).

56. Liang LW, Fifer MA, Hasegawa K, Maurer MS, Reilly MP & Shimada YJ. Prediction of genotype positivity in patients with hypertrophic cardiomyopathy using machine learning. Circ Genom Precis Med 14, e003259. doi:10.1161/circgen.120.003259 (2021).

