## Supplementary Material for "A genotype-phenotype taxonomy of hypertrophic cardiomyopathy"

### Supplemental Information

**Lara Curran[1,2 †], Antonio de Marvao[3,4,5 †], Paolo Inglese[3 †], Kathryn A McGurk[1,3], Pierre-Raphaël Schiratti[3], Adam Clement[3], Sean L Zheng[1,3], Surui Li[3,6], Chee Jian Pua[7], Mit Shah[3], Mina Jafari[3], Pantazis Theotokis[1,3], Rachel J Buchan[1,2,3], Sean J Jurgens[12,13], Claire E Raphael[1,2,11], Arun John Baksi[1,2], Antonis Pantazis[1,2], Brian P Halliday[1,2], Dudley J Pennell[1,2], Wenjia Bai[6,7], Calvin W L Chin[8,9,10], Rafik Tadros[12,14,15], Connie R Bezzina[12], Hugh Watkins[16,17], Stuart A Cook[3,8], Sanjay K Prasad[1,2], James S Ware[1,2,3], Declan P O'Regan[3 ✉]**

[1]National Heart and Lung Institute, Imperial College London, London, UK; [2]Royal Brompton & Harefield Hospitals, Guy's and St. Thomas' NHS Foundation Trust, London, UK; [3]MRC London Institute of Medical Sciences, Imperial College London, Hammersmith Hospital Campus, London, UK; [4]Department of Women and Children's Health, King's College London, London, UK; [5]British Heart Foundation Centre of Research Excellence, School of Cardiovascular Medicine and Sciences, King's College London, London, UK; [6]Biomedical Image Analysis Group, Department of Computing, Imperial College London, UK; [7]Department of Brain Sciences, Imperial College London, UK; [8]National Heart Research Institute Singapore, National Heart Center Singapore, Singapore; [9]Department of Cardiology, National Heart Center Singapore, Singapore; [10]Cardiovascular Sciences ACP, Duke NUS Medical School, Singapore; [11]Mayo Clinic Rochester, Minnesota, USA; [12]Department of Experimental Cardiology, University of Amsterdam, Amsterdam, Netherlands; [13]Cardiovascular Disease Initiative, Broad Institute of MIT and Harvard, Cambridge, MA, USA; [14]Cardiovascular Genetics Centre, Montreal Heart Institute, Montreal, QC, Canada; [15]Faculty of Medicine, Université de Montréal, Montreal, QC, Canada; [16]Radcliffe Department of Medicine, University of Oxford, Oxford, UK; [17]Wellcome Centre for Human Genetics, University of Oxford, Oxford, UK

### Estimation of wall thickness from meshes

Three dimensional meshes with the local wall thicknesses (WT) were generated from the myocardial segmentations of the end systolic (ES) and end diastolic (ED) phases, using previously published methods.[1–3] In order to make the modelling of the WT computationally tractable, the meshes were decimated by 99%. Specifically, because of the one-to-one correspondence between subjects' meshes vertices and atlas vertices, the decimation was applied to the atlas only, and the closest resulting vertices to the original atlas were selected from all meshes. This allowed us to preserve the correspondence between the individual vertices across all meshes.

### Unsupervised analysis of wall thickness values

Wall thicknesses were firstly adjusted by age at cardiac magnetic resonance (CMR) imaging, sex and ancestry using a linear regression, and normalized afterwards using the `Seurat` package for R.[4] To remove the intrinsic correlations between the values, due to the spatial nature of the data, and to reduce the effect of statistical noise, the data was compressed applying a principal components analysis (PCA), and retrieving the first 50 principal components.

A shared nearest neighbor graph (SNN) was built from the compressed data, where the nodes corresponded to the subjects, and the edges corresponded to the Jaccard index between the nearest neighbours of each pair of subjects. The nearest neighbors corresponded to the 20 subjects with the smallest cosine distance from each subject. Thus, the SNN graph was partitioned using the multilevel Louvain algorithm, with resolutions varying from 0.1 to 1 in steps of 0.1. Both SNN and its partitioning were performed using `Seurat` package for R.[4]

The optimal resolution was chosen by inspecting the cluster stability with `clustree`, corresponding to the partitioning before any mixing of cluster assignment was visible[5] (Supplementary Figs. 2, 3). Additionally, stability of the clusters was assessed by rerunning the clustering on 1000 random subsets, each with a size equal to 80% of the whole cohort, using the function `clusterboot` from the `fpc` package for R.[6]

If more than one partitioning was found corresponding to the same branch structure of the `clustree` plot, that with the greater stability was chosen (Supplementary Tables 1, 2).

### DDTree modelling of wall thickness values

DDRTree was applied on the adjusted wall thicknesses for age at CMR, sex and ancestry. Following the same approach described in the previous section, the first 50 principal components were calculated and used as input for the DDRTree algorithm.[7] All parameters were kept as default. The underlying tree structure, result of the procedure, was automatically partitioned into branches by considering as branching those points with a degree equal to 3. Subsequently, we manually merged small branches into their larger neighbour, such that the main structure of the tree was preserved. This step aimed at reducing the number of phenotypic sub-types and increase the statistical power of the subsequent modelling.

A set of clinical and anthropometric measures were tested for statistical association with the tree branches. We first applied a Kruskal Wallis test to continuous variables and $\chi^2$ test to discrete variables. Those variables showing a significant association (Benjamini-Hochberg adjusted $P < 0.05$) were post-hoc tested. A Dunn test was used to test each pair of branches, whereas the exact Fisher test was used for discrete variables in one-vs-all fashion. All *P*-values were adjusted for multiple testing using Benjamini-Hochberg method.

### External validation

Wall thicknesses were residualized by linear regression using sex and age at scan as covariates. In this way, we could compare the similarities between the intra-cohort statistical properties of WT values. The first 5 principal components were estimated from the development cohort and used to project the adjusted Singaporean WT values. The development cohort principal component scores were used to fit two random forest models aimed at predicting the x and y tree coordinates.

Performances of the models were evaluated using a 10-fold cross-validation, repeated 3 times. The fitted models were therefore used to predict the tree coordinates of the Singaporean individuals, from their principal component scores. Faithfulness of the tree mapping for the Singaporean cohort was evaluated by the "trustworthiness" $M_1$ measure, which estimates how observations that are similar in the original high dimensional space are placed close to each other in the low dimensional space. It ranges from 0 to 1, with larger values corresponding to a better representative low dimensional mapping.[8]

In order to evaluate the consistency of the local statistical patterns between the development tree and the projected Singaporean individuals, we considered Spearman's correlation between nearest neighbour points in the tree. We estimated the correlations between the adjusted WT of the closest points in the development tree, and compared their distribution with that of the correlations between the Singaporean WT and their closest points in the development tree. In order to test differences in the distributions, we used a Wilcoxon test for difference of medians, and a Kolmogorov Smirnov test for difference of distributions.

**Supplementary Table 1. Cluster stability at end diastole.** Results from the analysis of the cluster stability for end diastolic wall thickness (ED WT) in 1000 subsets. The possible values range between 0 and 1, where 0 means that the cluster is not stable, and 1 that the clusters are identical in all repetitions. Between a resolution of 0.4 and 0.5, determined from the `clustree` plot (Supplementary Fig. 2). We chose the resolution of 0.5 because it is also characterized by more stable clusters. Cluster 2 is found as an intermediate state between cluster 0 and 1, and it is less stable than the other two.

| resolution | cluster 0 | cluster 1 | cluster 2 |
|---|---|---|---|
| 0.4 | 0.95 | 0.85 | 0.50 |
| **0.5** | **0.95** | **0.92** | **0.78** |

**Supplementary Table 2. Cluster stability at end systole.** Results from the analysis of the cluster stability for end systolic wall thickness (ES WT) WT in 1000 subsets. The possible values range between 0 and 1, where 0 means that the cluster is not stable, and 1 that the clusters are identical in all repetitions. Resolutions from 0.1 to 0.6 are determined from the `clustree` plot (Supplementary Fig. 3). The resolution of 0.1 was chosen because it is the lowest value among those with more stable clusters.

| resolution | cluster 0 | cluster 1 |
|---|---|---|
| **0.1** | **0.96** | **0.96** |
| 0.2 | 0.96 | 0.96 |
| 0.3 | 0.96 | 0.96 |
| 0.4 | 0.96 | 0.96 |
| 0.5 | 0.96 | 0.96 |
| 0.6 | 0.94 | 0.95 |

**Supplementary Table 3. Pathogenic/likely pathogenic variant prediction from tree coordinates.** Fitted parameters for the GAM model used to predict individuals with P/LP variants using the 2 tree coordinates. [1]OR, Odds ratio; CI, Confidence interval

| | ED | | | ES | | |
|---|---|---|---|---|---|---|
| **Characteristic** | **log(OR)[1]** | **95% CI[1]** | **p-value** | **log(OR)[1]** | **95% CI[1]** | **p-value** |
| (Intercept) | -1.1 | -1.4, -0.93 | <0.001 | -1.2 | -1.4, -0.96 | <0.001 |
| s(Z1) | | | 0.008 | | | 0.013 |
| s(Z2) | | | 0.2 | | | 0.062 |

**Supplementary Table 4. Singaporean branch assignment.** Singaporean HCM patients were assigned to the tree branches of their nearest neighbours in the development tree. In both end diastole (ED) and end systole (ES), most of the individuals were assigned to branches 1 and 4.

| Branch | ED | ES |
|---|---|---|
| 1 | 23 | 21 |
| 2 | 13 | 1 |
| 3 | 4 | 2 |
| 4 | 20 | 35 |
| 5 | - | 1 |

### Dimensionality reduction and unsupervised clustering of clinical features

Participant features comprised demographic data, clinical characteristics, CMR and echocardiographic measurements, and reported interventions and medicines (Supplementary Table 9). Missing values were inferred with the `mice` package for R.[9] Numerical features were converted to categorical variables by clustering groups of values into bins with a K-means algorithm.[10] All categorical variables were then transformed into binary variables with one-hot encoding.

Dimensionality reduction was performed on this collection of binary variables with UMAP (uniform manifold approximation and projection)[11] using the following parameters: Dice metric, 25 components, 8 neighbouring sample points and a minimum distance between points of $10^{-6}$. Finally, unsupervised clustering was applied to the 25 resulting UMAP components with a K-means algorithm, revealing three clusters (Supplementary Fig. 8A). Genotype status was found to be significantly associated with the clusters, using a $\chi^2$ test. A post-hoc exact Fisher test was then performed to find cluster-specific enrichment. Adjustment for multiple testing was done with the Benjamini-Hochberg procedure, $P < 0.05$. Cluster 1 was significantly enriched with genotype-negative (NEG) subjects while cluster 3 was associated with genotype-positive (P/LP) and genotype-indeterminate (VUS) individuals (Supplementary Fig. 8C). Feature importance from the initial set of participant data was assessed by applying a Kruskal-Wallis test for numerical features and a $\chi^2$ test for categorical features. Significant associations (adjusted with the Benjamini-Hochberg method, $P < 0.05$) were further tested for cluster-specific enrichment: a Dunn test was used to test each pair of numerical features (Supplementary Fig. 8D), while an exact Fisher test looked for one-vs-rest differences in categorical features (Supplementary Fig. 8E).

The clustering revealed features characterising each group: 1) older female participants with lower body surface area (BSA), lower left ventricular (LV) volume, higher ejection fraction, hypertension, low activity score and on beta blockers and diuretic medications; 2) male participants with higher BSA, higher LV mass, higher LV maximum wall thickness, hypertension, moderate activity score and on medications for blood pressure (ACE/ARBs) and protective vascular (ASA/Clopi) medications; 3) younger participants with a family history of hypertrophic cardiomyopathy (HCM), no clinical cardiovascular risk factor and a high activity score.

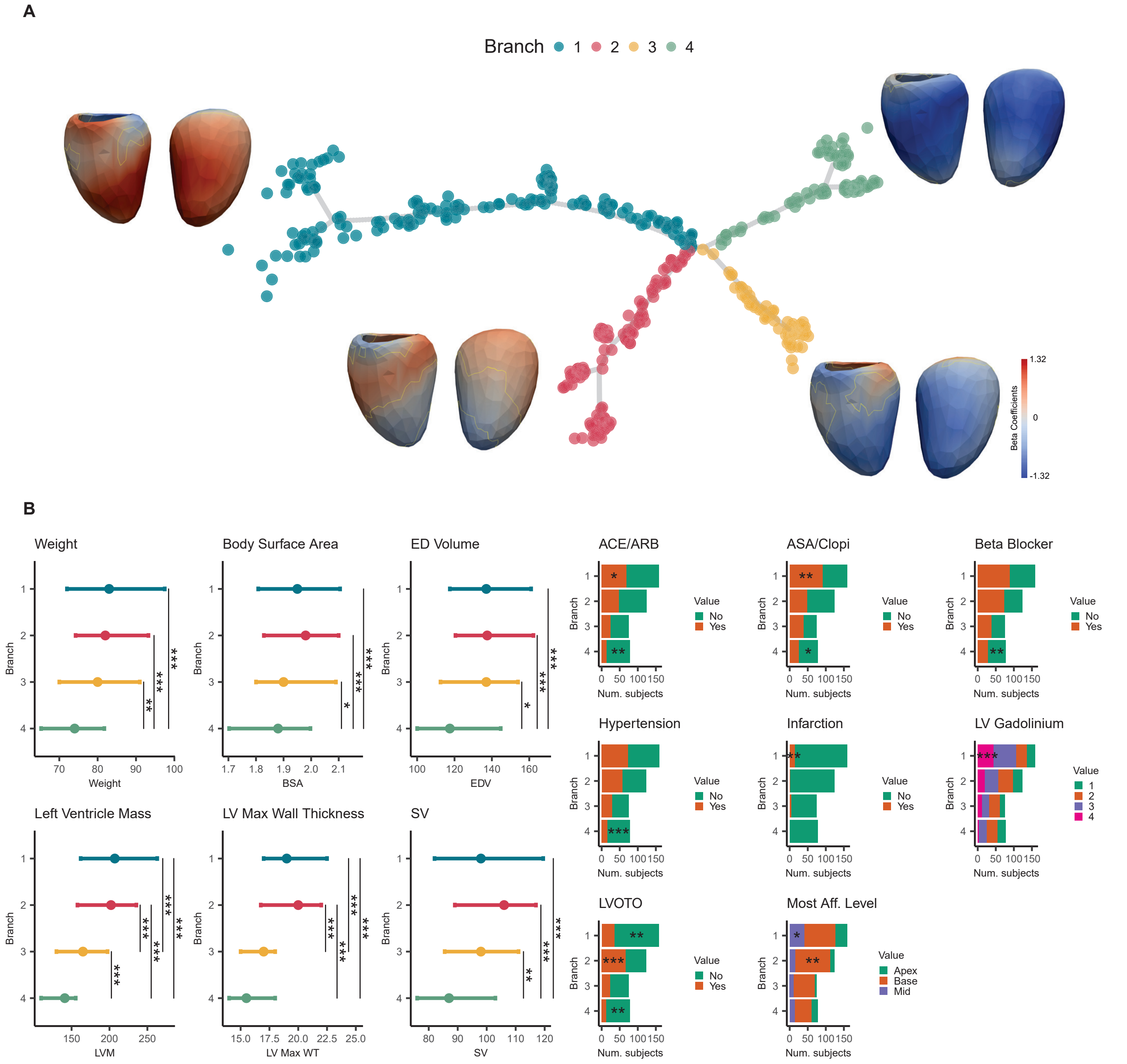


**Supplementary Figure 1. Phenotypic tree from 3D end-diastolic wall thickness. a.** The projection of patients' 3D end-diastolic (ED) wall thickness (WT) by the DDRTree dimensionality reduction reveal the presence of four main branches that are associated to specific morphological changes of the myocardium. Each branch is represented by the decimated ED atlas mesh, coloured accordingly to the beta coefficients resulting from testing the average difference between each branch individual and the other subjects. The yellow contour denotes the areas with a beta significantly different from zero. **b.** The continuous and discrete phenotypic variables found to be significantly associated to at least one branch. For left ventricular (LV) Gadolinium, labels are as follows: 1: None, 2: Minimal, 3: Moderate and 4: Severe. The significance for the enrichment of discrete variables is reported within the bars. ACE, Angiotensin-converting enzyme inhibitors; Aff, affected; ARB, Angiotensin receptor blockers; ASA, aspirin; Clopi, clopidogrel; LVOTO, Left ventricular outflow tract obstruction; SV, stroke volume. Only the significant pairs are reported with the symbols: $* \ P \leq 0.05; ** \ P \leq 0.01; *** \ P \leq 0.001; **** \ P \leq 0.0001, n = 436.$

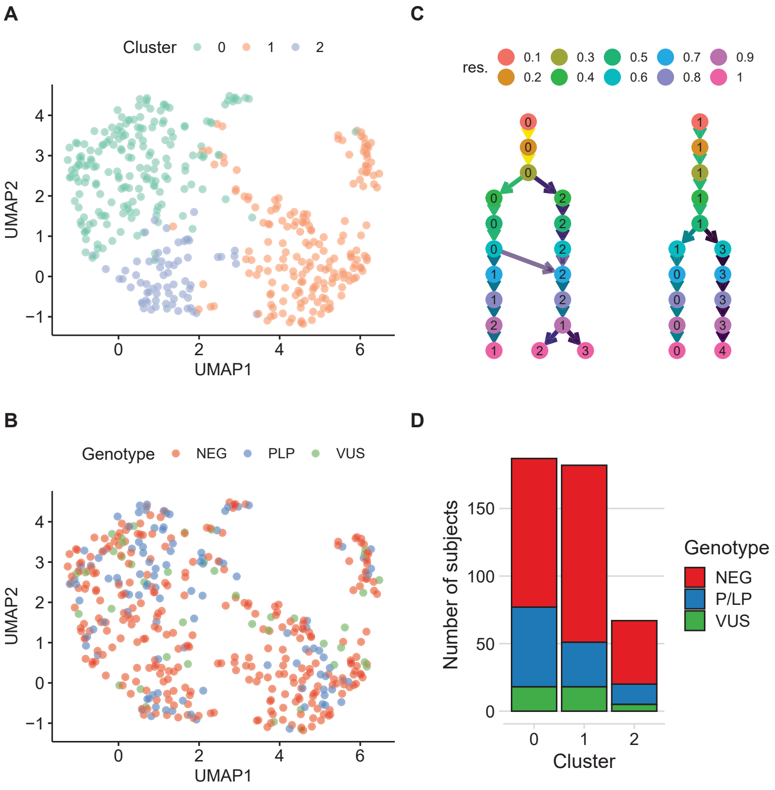


**Supplementary Figure 2. Selection of optimal resolution for clustering of end diastolic wall thickness.** The optimal resolution for the Louvain partitioning is found by inspecting the `clustree` plot (**c.**). In this case, the value of 0.5 was chosen, corresponding to the resolution with stable branching before any assignment mixing (diagonal arrows), and the largest bootstrapping stability (Supplementary Table 1). The UMAP projections in **a.** and **b.** show the parallelism between the clusters and the genotypes. **d.** Genotype proportions by cluster.

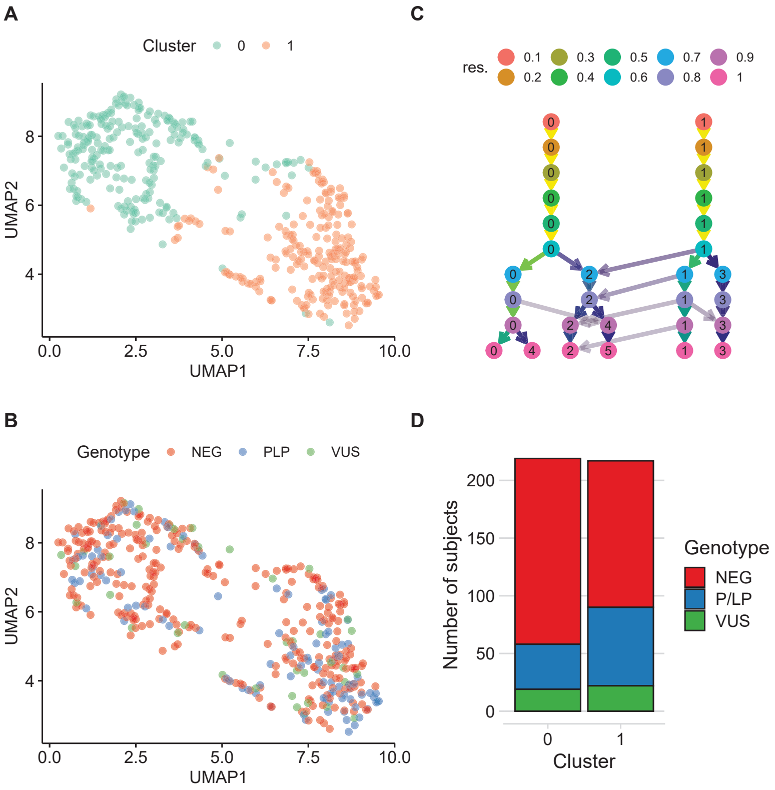


**Supplementary Figure 3. Selection of optimal resolution for clustering of end systolic wall thickness.** The optimal resolution for the Louvain partitioning is found by inspecting the `clustree` plot (**c.**). In this case, the value of 0.1 was chosen, corresponding to the resolution with stable branching before any assignment mixing (diagonal arrows), and the largest bootstrapping stability (Supplementary Table 2). The UMAP projections in **a.** and **b.** show the parallelism between the clusters and the genotypes. **d.** Genotype proportions by cluster.

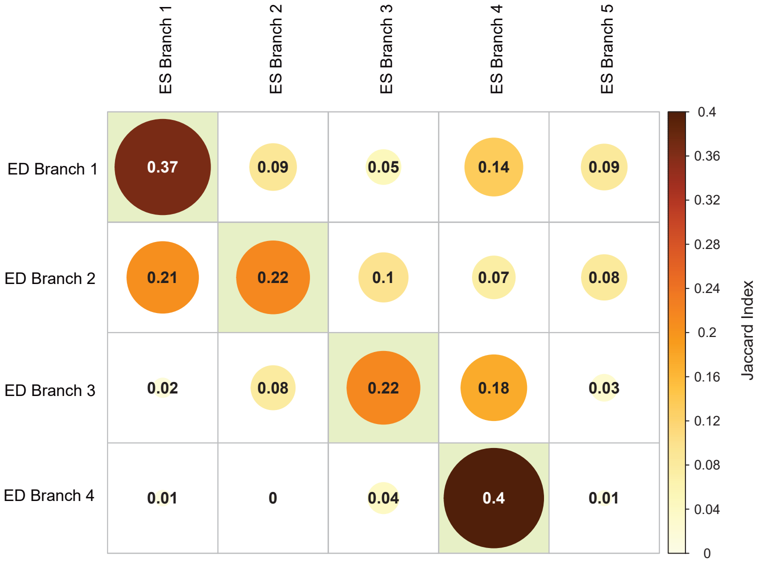


**Supplementary Figure 4. Co-occurrence between end diastolic and end systolic tree branches.** The Jaccard index of the subjects membership for the branches in the DDRTree from ED and ES WT shows that branches 1 to 4 have the largest co-occurrence and they can be considered capturing a similar phenotypic subpopulation. Branch 5 in end systolic DDRTree is not found in the end diastolic DDRTree and consists of an average sub-type of the cohort.

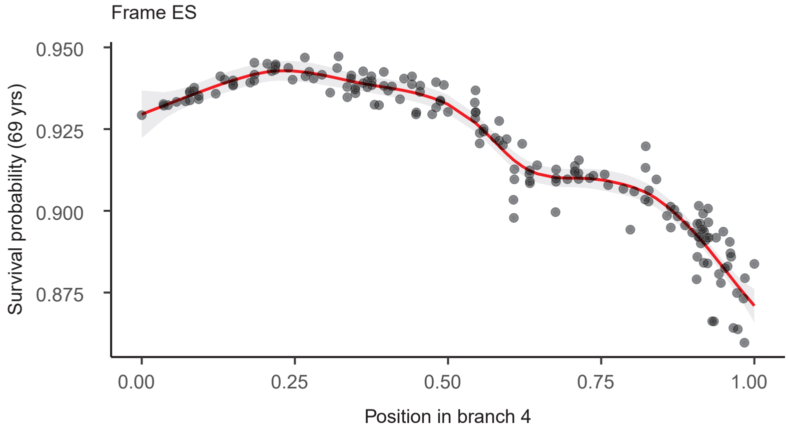


**Supplementary Figure 5. Survival probability in end systolic branch 4.** The more distal regions of branch 4 correspond to lower probability of survival at a chronological age of 69 years. The OR between the distal and base points of the branch is 0.9773.

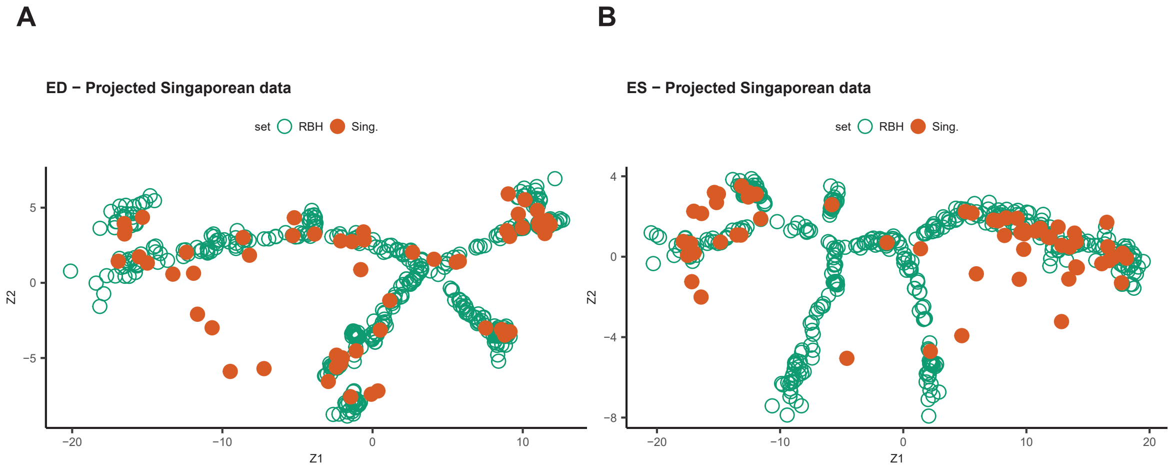


**Supplementary Figure 6. Predicted tree coordinates for the Singaporean cohort.** The coordinates predicted by the two random forest models for the Singaporean cohort follow the original spatial distribution of the development cohort, with few points falling outside the main structure of the tree, in both end diastole (**a.**) and end systole (**b.**). Sing, Singaporean patients; RBH, Royal Brompton Hospital patients.

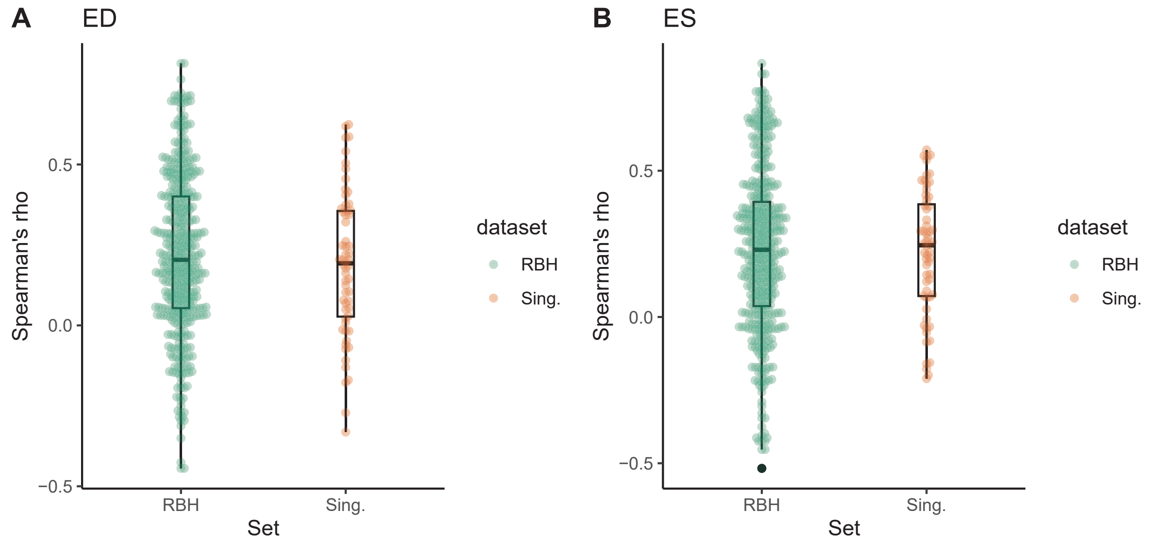


**Supplementary Figure 7. Similarity between nearest tree points.** Spearman's correlation between the adjusted wall thickness of the nearest RBH points in the tree follow the same distribution of the nearest Singaporean and RBH points, in both end diastole (**a.**) and end systole (**b.**). Sing, Singaporean patients; RBH, Royal Brompton Hospital patients.

**Supplementary Table 5. Participant characteristics and CMR-derived cardiac measurements in UK Biobank.** BSA, body surface area; concentricity, (left ventricular mass / left ventricular end-diastolic volume); CMR, cardiac magnetic resonance imaging; DBP, diastolic blood pressure; EDV, end-diastolic volume; EF, ejection fraction; ESV, end-systolic volume; FD, fractal dimension; LV, left ventricular; LVM, left ventricular mass; LVMI, left ventricular mass index (LVM/BMI); peak diastolic strain rate, PDSR; RA, right atrial; RV, right ventricular; SBP, systolic blood pressure; WT, wall thickness. *Medication for cholesterol, blood pressure, diabetes.

| **Characteristic** | **UKBB n=16,691** |
|---|---|
| Female | 8,775 (52.5) |
| Age at scan, y | 55 ± 7.5 |
| White | 14,683 (87.9) |
| BSA, $m^2$ | 1.9 ± 0.2 |
| LVEDV, ml | 148 ± 33.5 |
| LVESV, ml | 60.4 ± 19 |
| LVEF, ml | 59.6 ± 6 |
| LVM, g | 86 ± 22.1 |
| LVMI, $g/m^2$ | 45.8 ± 8.5 |
| LV maximum WT, mm | 9.4 ± 1.6 |
| Mean apical FD | 1.21 ± 0.05 |
| Mean basal FD | 1.19 ± 0.03 |
| Mean global FD | 1.17 ± 0.03 |
| LV global radial strain, % | 45 ± 8.3 |
| LV global circumferential strain, % | -22.3 ± 3.4 |
| LV global longitudinal strain, % | -18.5 ± 2.8 |
| LV radial PDSR | -5.7 ± 2 |
| LV longitudinal PDSR | 1.7 ± 0.6 |
| LV concentricity, g/mL | 0.58 ± 0.08 |
| Heart rate, min | 69.5 ± 11.6 |
| Hypertension | 4,857 (29) |
| On medication* | 2,241 (13.4) |
| SBP, mmHg | 137.5 ± 18.1 |
| DBP, mmHg | 78.6 ± 9.9 |

**Supplementary Table 6. Characteristics of Singaporean HCM cohort**. BSA, body surface area; SBP, systolic blood pressure.

| **Characteristic** | **Singaporean HCM n=60** |
|---|---|
| Female | 11 (11.7) |
| Age at scan, y | 58.9 ± 20 |
| Chinese | 52 (86.7) |
| BSA, $m^2$ | 1.8 ± 0.2 |
| SBP, mm Hg | 137 ± 24.8 |
| Genotype SARC-NEG | 28 (46.7) |
| Genotype SARC-VUS | 16 (26.7) |
| Genotype SARC-P/LP | 16 (26.7) |

**Supplementary Table 7. Cumulative hazard model.** All cause mortality in individuals with hypertrophic cardiomyopathy carrying pathogenic or likely pathogenic sarcomeric variants (SARC-P/LP) compared to those without variants in genes that may cause or mimic HCM (SARC-NEG) and those with variants of uncertain significance (SARC-VUS), adjusted for Age, Sex and Race. n = 436; $P$ = 0.002. [1]HR = Hazard Ratio, CI = Confidence Interval

| | **Full model** | | | **Genotype only** | | |
|---|---|---|---|---|---|---|
| **Characteristic** | **HR[1]** | **95% CI[1]** | **p-value** | **HR[1]** | **95% CI[1]** | **p-value** |
| **P/LP** | | | | | | |
| N | — | — | | — | — | |
| Y | 2.63 | 1.43, 4.86 | 0.002 | 2.62 | 1.42, 4.84 | 0.002 |
| **Race** | | | | | | |
| White | — | — | | | | |
| Other | 0.68 | 0.34, 1.37 | 0.3 | | | |
| **Sex** | | | | | | |
| F | — | — | | | | |
| M | 1.15 | 0.72, 1.85 | 0.6 | | | |

**Supplementary Table 8. Cumulative hazard model excluding SARC-VUS.** All cause mortality in individuals with hypertrophic cardiomyopathy carrying pathogenic or likely pathogenic sarcomeric variants (SARC-P/LP) compared to those without variants in genes that may cause or mimic HCM (SARC-NEG), adjusted for Age, Sex and Race. N = 395; P = 0.003. [1]HR = Hazard Ratio, CI = Confidence Interval

| | Full model | | | Genotype only | | |
|---|---|---|---|---|---|---|
| **Characteristic** | **HR**[1] | **95% CI**[1] | **p-value** | **HR**[1] | **95% CI**[1] | **p-value** |
| **P/LP** | | | | | | |
| N | — | — | | — | — | |
| Y | 2.66 | 1.43, 4.95 | 0.002 | 2.64 | 1.40, 4.98 | 0.003 |
| **Race** | | | | | | |
| White | — | — | | | | |
| Other | 0.68 | 0.30, 1.54 | 0.4 | | | |
| **Sex** | | | | | | |
| F | — | — | | | | |
| M | 1.26 | 0.77, 2.06 | 0.4 | | | |

**Supplementary Table 9. Participant clinical features.** MRI, magnetic resonance imaging; HCM, hypertrophic cardiomyopathy; SCD, sudden cardiac death CCS, Canadian Cardiovascular Society[12]; NYHA, New York Heart Association[13]; LV, left ventricular; RV, right ventricular; ACE, angiotensin-converting enzyme; ARB, angiotensin receptor blockers; ASA, acetylsalicylic acid.

| **Demographics** | **Mean ± SD or n (%)** |
|---|---|
| Age at time of MRI scan (years) | 57.3 (± 14.4) |
| Sex, male | 310 (71.1%) |
| Ethnicity | |
| non-Finnish European | 353 (81%) |
| South Asian | 52 (11.9%) |
| African | 16 (3.7 %) |
| Others | 14 (3.2 %) |
| East Asian | 1 (0.2 %) |
| **Clinical characteristics** | |
| Body surface area ($m^2$) | 2 (± 0.3) |
| Diastolic blood pressure (mmHg) | 76.2 (± 11.4) |
| Systolic blood pressure (mmHg) | 133 (± 18.5) |
| Pulse rate (bpm) | 70.1 (± 13.7) |
| Smoker | 172 (39.4%) |
| Alcohol intake (units per week) | 6.8 (± 12.3) |
| Activity score | |
| 0 | 64 (14.7%) |
| 1 | 68 (15.6%) |
| 2 | 252 (57.8%) |
| 3 | 49 (11.2%) |
| 4 | 3 (0.7%) |
| Hypertension | 175 (40.1%) |
| Diabetes mellitus | 47 (10.8%) |
| Coronary artery disease | 46 (10.6%) |
| Myocardial infarction | 21 (4.8%) |
| Family history of HCM | 85 (19.5%) |
| Family history of SCD | 70 (16.1%) |
| CCS Angina Grading Scale | |
| 0 | 57 (13.1%) |
| I | 277 (63.5%) |
| II | 88 (20.2%) |
| III | 13 (3.0%) |
| IV | 1 (0.2%) |
| NYHA Classification of Heart Failure | |
| No heart failure | 48 (11.0%) |
| I | 177 (40.6%) |
| II | 176 (40.4%) |
| III | 31 (7.1%) |
| IV | 4 (0.9%) |

| **Measurements derived from MRI and echocardiogram** | **Mean ± SD or n (%)** |
|---|---|
| LV end-diastolic volume (mL) | 137.6 (± 34.5) |
| LV end-systolic volume (mL) | 37.3 (± 18.3) |
| LV stroke volume (mL) | 100 (± 24.4) |
| LV ejection fraction | 73.5 (± 8.5) |
| LV mass (mL) | 189 (± 67) |
| LV maximum wall thickness (mm) | 18.8 (± 4.5) |
| LV most affected segment | |
| Anterior | 43 (12.0%) |
| Inferior | 8 (2.2%) |
| Lateral | 11 (3.1%) |
| Septal | 295 (82.6%) |
| LV most affected level | |
| Base | 210 (59.3%) |
| Mid | 78 (22.0%) |
| Apex | 66 (18.6%) |
| Mitral regurgitation | |
| None | 203 (54.9%) |
| Minimal | 124 (33.5%) |
| Moderate | 40 (10.8%) |
| Severe | 3 (0.8%) |
| LV gadolinium | |
| None | 53 (14.0%) |
| Minimal | 124 (32.7%) |
| Moderate | 133 (35.1%) |
| Severe | 69 (18.2%) |
| RV hypertrophy | 45 (10.3%) |
| Coincident infarction | 19 (4.4%) |
| LV outflow tract obstruction | 120 (27.5%) |
| LV outflow peak velocity (m/s) | 2.3 (± 0.7) |
| **Interventions and medicines** | |
| ACE inhibitors and ARBs | 145 (33.3%) |
| ASA/clopidrogel | 184 (42.2%) |
| Beta blocker | 210 (48.2%) |
| Diuretic | 66 (15.1%) |

**Supplementary Figure 8. Unsupervised clustering of clinical features and feature importance. a.** Participant clinical features (Supplementary Table 9) segmented in three clusters with a K-means algorithm projected in the 2-dimensional space of the first two UMAP components. **b.** Genotype status of participants in the 2-D UMAP space. **c.** Distribution of genotype status by cluster. **d.** Significant pairs of associations between identified clusters and numerical features from the initial set of data. The line represents the interquartile range and median value. **e.** Significant one-vs-rest associations between clusters and categorical features from the initial set of data grouped by significance level. The height of the curved bars illustrates the significance level ($-log_{10}P$). Only the significant pairs are reported with the symbols: $*\ P \leq 0.05; **\ P \leq 0.01; ***\ P \leq 0.001; ****\ P \leq 0.0001, n = 436.$

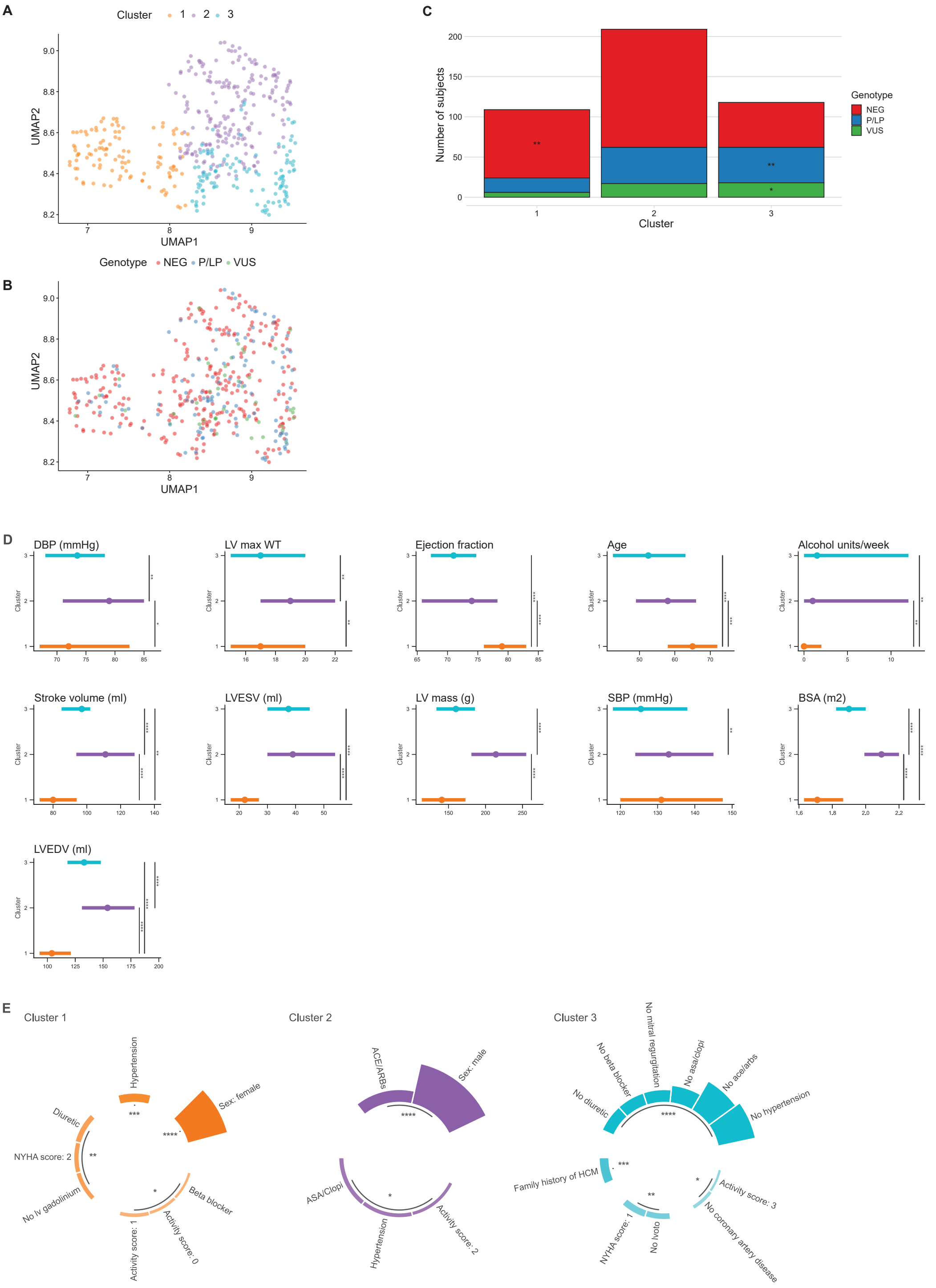

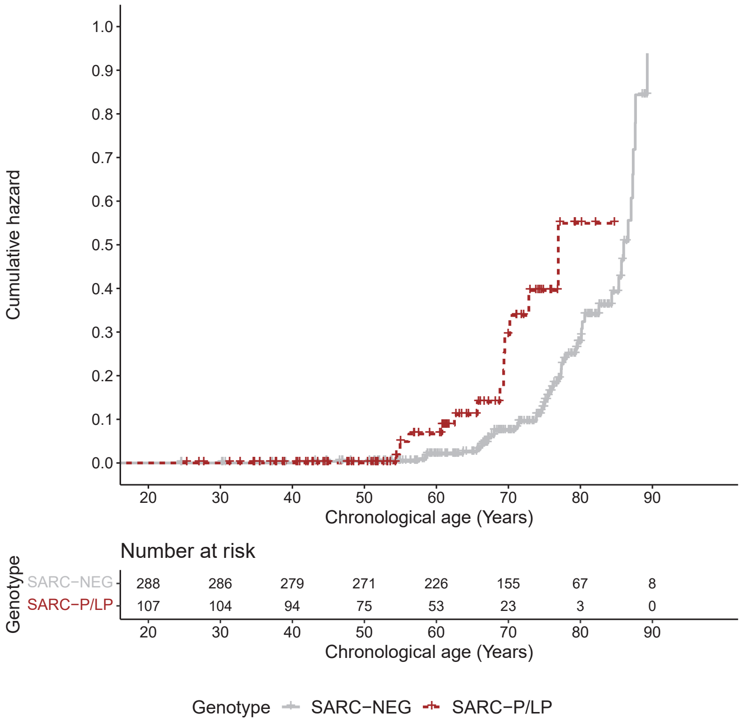


**Supplementary Figure 9. Cumulative hazard plot.** All cause mortality in individuals with hypertrophic cardiomyopathy (HCM) carrying pathogenic or likely pathogenic sarcomeric variants (SARC-P/LP) compared to those without variants in genes that may cause or mimic HCM (SARC-NEG) (Hazard ratio 2.66; 95% CI: 1.42-4.96; *P* = 0.002).

### Variant curation pipeline

This pipeline can be found on GitHub (https://github.com/ImperialCollegeLondon/HCM-taxonomy) and has been previously published.[14] All genetic data was annotated using Ensembl Variant Effect Predictor (VEP; version 105)[15] with plugins for NMD, SpliceAI (version 1.3.1),[16] ClinVar (version 2022 01 15),[17] gnomAD (version r2.1),[18] and LOFTEE.[18] The VEP output was analysed using R (version 3.6.0).

Protein-altering variants, defined using MANE transcripts, that had a MAF of <0.1% in gnomAD for variants identified in cases, and <0.1% in gnomAD and UK Biobank for variants identified in the UK Biobank, were included in the analyses. Protein altering variants were specified as high or moderate impact by Sequence Ontology and Ensembl, with the addition of splice region variants for further curation. The variants were filtered for genes and protein consequences of interest,[19] to include 8 definitive-evidence sarcomeric HCM genes (*MYH7, MYBPC3, MYL2, MYL3, ACTC1, TNNI3, TNNT2, TPM1*), 3 medium-evidence HCM genes (*CSRP3, TNNC1, JPH2*), 2 intrinsic cardiomyopathy genes (*ACTN2* (moderate classification), *PLN* (definitive classification)), and 12 syndromic genes that can cause isolated left ventricular hypertrophy (*FHL1, TTR, FLNC, GLA, LAMP2, PRKAG2, PTPN11, RAF1, RIT1, ALPK3, CACNA1C, DES*). *FLNC, ALPK3, ABCC9, CRYAB, MYO6, and RIT1*, were not sequenced in cases, but were analysed in UK biobank. No protein-altering variants were identified in *TNNC1* in the case cohort.

Splice region variants (outside the canonical splice donor and acceptor sites) were assessed in two ways; i) via ClinVar report: splice region variants found pathogenic with at least 2 star evidence for HCM in ClinVar and reported functional evidence for splicing were termed "splice confirmed"; if the functional evidence was unclear the protein consequence remained unchanged; if there was functional evidence of an alternative mechanism to splicing, the protein consequence was renamed (e.g. missense variant); ii) via prediction threshold: of the splice region variants, they were excluded if they did not meet the spliceAI threshold of >0.8, and these thresholds were used to identify potentially splice-causing variants of those splice region variants identified with a non-synonymous consequence flag (e.g. intron variant).

The pipeline then consisted of three main filtering steps which resulted in an output of four columns of binary code flagging genotype status (heterozygous, compound heterozygotes, and homozygotes, combined) as "1": SARC-NEG – Individuals who do not harbor any rare non-synonymous variants in any of the 25 genes of interest. This was a stringent filter to identify an unambiguous genotype-negative control group.

SARC-VUS – Individuals harboring rare variants in one or more of the 8 definitive HCM-associated sarcomere-encoding genes. Rare variants were restricted to known disease-associated variant classes. This step separated the variants into two subsets: i) Loss of function (LoF) alleles (group A), which contained only the gene *MYBPC3*, and filters for the protein consequences of stop gained, splice acceptor variant, splice donor variant, frameshift variant, and splice region variant (with additional in silico evidence of an effect on splicing). LOFTEE was incorporated in this step to exclude loss of function (LoF) variants that were flagged as "low confidence" (LC) and other LOFTEE flags, such as "NAGNAG sit" requiring reannotation to non-LoF variant status; ii) Protein altering (PAV) alleles (group B), which included all 8 sarcomeric genes, including *MYBPC3*, and filters for the protein consequences of missense variant, inframe insertion, and inframe deletion.

Both groups included additional positional annotation (LoF variants found in the last exon or 55bp into the penultimate exon or stop gained variants with a NMD flag using the NMD plugin), this included variants that introduce a protein-truncating variant (PTC) and predicted to lead to nonsense-mediated decay (NMD). The variants flagged 'coding sequence variant' and 'protein altering variant' were manually curated, as were 'stop lost' and 'start lost' which were examined via ENSEMBL sequence and UCSC Genome Browser to identify in-frame rescues nearby. To be included in the SARC VUS group, the variants were required to meet a maximum gnomAD filter allele frequency (FAF) threshold for HCM (<0.00004) and excluded variants deemed P/LP for DCM on ClinVar.

SARC-P/LP is as SARC-VUS, plus annotated as P/LP according to the ACMG guidelines.[20] Variants were reviewed if reported as P/LP for HCM by at least one submitter in ClinVar, or if flagged as P/LP by the CardioClassifier decision support software.[21] Variants that did not meet either of these criteria were not individually reviewed.

### References


1. Biffi C, de Marvao A, Attard MI, Dawes TJW, Whiffin N, Bai W, Shi W, Francis C, Meyer H, Buchan R, Cook SA, Rueckert D & O'Regan DP. Three-dimensional cardiovascular imaging-genetics: a mass univariate framework. *Bioinformatics* **34,** 97–103. doi:10.1093/bioinformatics/btx552 (2018).
2. Bhuva AN, Treibel TA, De Marvao A, Biffi C, Dawes TJW, Doumou G, Bai W, Patel K, Boubertakh R, Rueckert D, O'Regan DP, Hughes AD, Moon JC & Manisty CH. Sex and regional differences in myocardial plasticity in aortic stenosis are revealed by 3D model machine learning. *Eur Heart J Cardiovasc Imaging* **21,** 417–427. doi:10.1093/ehjci/jez166 (2020).
3. Schafer S *et al.* Titin-truncating variants affect heart function in disease cohorts and the general population. *Nat Genet* **49,** 46–53. doi:10.1038/ng.3719 (2017).
4. Hao Y *et al.* Integrated analysis of multimodal single-cell data. *Cell* **184,** 3573–3587.e29. doi:10.1016/j.cell.2021.04.048 (2021).
5. Zappia L & Oshlack A. Clustering trees: a visualization for evaluating clusterings at multiple resolutions. *GigaScience* **7.** doi:10.1093/gigascience/giy083 (2018).
6. Hennig C. *fpc: Flexible Procedures for Clustering* R package version 2.2-9 (2020). https://CRAN.R-project.org/package=fpc.
7. Qiu X, Mao Q, Tang Y, Wang L, Chawla R, Pliner HA & Trapnell C. Reversed graph embedding resolves complex single-cell trajectories. *Nat Methods* **14,** 979–982. doi:10.1038/nmeth.4402 (2017).
8. Van der Maaten L. *Learning a Parametric Embedding by Preserving Local Structure* in *Proceedings of the Twelfth International Conference on Artificial Intelligence and Statistics* (eds van Dyk D & Welling M) **5** (PMLR, Hilton Clearwater Beach Resort, Clearwater Beach, Florida USA, 2009), 384–391.
9. Van Buuren S & Groothuis-Oudshoorn K. mice: Multivariate Imputation by Chained Equations in R. *J Stat Softw* **45,** 1–67. doi:10.18637/jss.v045.i03 (2011).
10. Lloyd SP. Least-Squares Quantization in PCM. *IEEE Trans Inf Theory* **28,** 129–137. doi:10.1109/Tit.1982.1056489 (1982).
11. McInnes L, Healy J, Saul N & Großberger L. UMAP: Uniform Manifold Approximation and Projection. *J Open Source Softw* **3,** 861. doi:10.21105/joss.00861 (2018).
12. Campeau L. Grading of Angina-Pectoris. *Circulation* **54,** 522–523. doi:10.1161/circ.54.3.947585 (1976).
13. Dolgin M. *Nomenclature and criteria for diagnosis of diseases of the heart and great vessels* 9th / editor, Martin Dolgin / associate editors, Arthur C. Fox, Richard Gorlin, Richard I. Levin / Criteria Committee, Richard P. Devereaux ... [et al.] doi:10.1001/jama.1940.02810200082036 (Little, Brown, Boston ; London, 1994).
14. De Marvao A *et al.* Phenotypic Expression and Outcomes in Individuals With Rare Genetic Variants of Hypertrophic Cardiomyopathy. *J Am Coll Cardiol* **78,** 1097–1110. doi:10.1016/j.jacc.2021.07.017 (2021).
15. McLaren W, Gil L, Hunt SE, Riat HS, Ritchie GR, Thormann A, Flicek P & Cunningham F. The Ensembl Variant Effect Predictor. *Genome Biol* **17,** 122. doi:10.1186/s13059-016-0974-4 (2016).
16. Jaganathan K, Kyriazopoulou Panagiotopoulou S, McRae JF, Darbandi SF, Knowles D, Li YI, Kosmicki JA, Arbelaez J, Cui W, Schwartz GB, Chow ED, Kanterakis E, Gao H, Kia A, Batzoglou S, Sanders SJ & Farh KK. Predicting Splicing from Primary Sequence with Deep Learning. *Cell* **176,** 535–548 e24. doi:10.1016/j.cell.2018.12.015 (2019).
17. Landrum MJ *et al.* ClinVar: improving access to variant interpretations and supporting evidence. *Nucleic Acids Res* **46,** D1062–D1067. doi:10.1093/nar/gkx1153 (2018).
18. Karczewski KJ *et al.* The mutational constraint spectrum quantified from variation in 141,456 humans. *Nature* **581,** 434–443. doi:10.1038/s41586-020-2308-7 (2020).
19. Ingles J *et al.* Evaluating the Clinical Validity of Hypertrophic Cardiomyopathy Genes. *Circ Genom Precis Med* **12,** e002460–e002460. doi:10.1161/CIRCGEN.119.002460 (2019).
20. Richards S, Aziz N, Bale S, Bick D, Das S, Gastier-Foster J, Grody WW, Hegde M, Lyon E, Spector E, Voelkerding K & Rehm HL. Standards and guidelines for the interpretation of sequence variants: a joint consensus recommendation of the American College of Medical Genetics and Genomics and the Association for Molecular Pathology. *Genet Med* **17,** 405–24. doi:10.1038/gim.2015.30 (2015).
21. Whiffin N *et al.* CardioClassifier: disease- and gene-specific computational decision support for clinical genome interpretation. *Genet Med* **20,** 1246–1254. doi:10.1038/gim.2017.258 (2018).
